# Pandemic waves as the outcome of coupled behaviour and disease dynamics: a mathematical modelling study

**DOI:** 10.64898/2026.02.05.26345658

**Authors:** Sefah Frimpong, Chris T. Bauch

## Abstract

**Background:** The COVID-19 pandemic was strongly shaped by the interaction between population behaviour and transmission dynamics. Standard mathematical models do not account for this interaction, however.

**Objective:** we tested whether adding a mechanistic representation of population behavioural dynamics improves the ability of a mathematical model to explain and predict COVID-19 pandemic waves.

**Methods:** We compared a standard Susceptible-Infected-Recovered (SIR) model to a variant (SIRx) with a mechanistic representation of behavioural processes, including two-way coupling between behaviour and transmission dynamics. We used approximate Bayesian computation to parameterise the models with SARS-CoV-2 case incidence and the Oxford stringency index from 13 European countries. Models were fitted to the Spring 2020 wave, and their out-of-sample prediction for the Summer/Fall 2020 wave was tested. Outcome measures included the Akaike Information Criterion (AICc), the area between empirical and model epidemic curves, and predicted timing/magnitude of the second wave.

**Results:** The average AICc for the SIRx model across all 13 countries was lower (−2638±345 versus − 2295±212 for SIR), meaning that the SIRx model explains the data more parsimoniously. The average area-between-curves was also lower (0.072±0.071 versus 0.16±0.11). The predicted peak magnitude for the SIRx model (0.0015±0.0014) was closer to the data (0.0006±0.0005) than the SIR prediction (0.0083±0.0090). The average day-of-peak for the SIRx model (283±19 days from first data point) was also closer to the data (278±25), than the SIR prediction (253±31), although the 95% credible intervals for individual countries were very large.

**Conclusion:** Coupling behavioural and disease dynamics improves the ability of mathematical models to explain and predict crucial features of pandemic waves.

**Research in context:** *Evidence before this study:* Most mathematical models of infectious disease transmission do not explicitly account for behaviour, but the COVID-19 pandemic clarified the role of behavioural processes in determining the trajectory of infectious diseases in populations. On the other hand, many theoretical models of coupled behaviour-disease processes exist, although relatively few attempt to validate these models against data. We searched Google Scholar using the terms COVID-19 model, and behavio*-disease or behavio* epidem* from March 1, 2020 to October 8, 2025. We did not find any papers that compared retrospective out-of-sample model predictions of COVID-19 pandemic waves of a non-behavioural transmission model to the predictions of a coupled behaviour-disease model, in multiple populations.

*Added value of this study:* We carried out such a comparison for 13 European countries, by fitting models to the first COVID-19 wave in Spring 2020 and testing how well they would have predicted the second wave. We found that the coupled behaviour-disease model predicted the second wave better than the non-behavioural model, and was also more parsimonious, despite having more parameters. This study shows that feedback between disease dynamics and behavioural dynamics is a significant factor for determining the timing and magnitude of pandemic waves caused by an acute respiratory infection. It also shows that integrating population behaviour dynamics into transmission models is feasible, and can better explain observed temporal patterns in case incidence.

*Implications of all the available evidence:* Mathematical models that endogenously include the feedback between infectious disease dynamics and behavioural dynamics can add a unique and complementary tool to the public health modelling toolbox during a pandemic. Such models could help design public health interventions by improving our ability to anticipate population responses to both the interventions themselves, and a rapidly evolving epidemiological landscape.

## Background

The coronavirus disease 2019 (COVID-19) pandemic impacted health and livelihoods around the world [1]. In the early stages of the pandemic, due to a lack of pharmaceutical interventions, the need for non-pharmaceutical interventions (NPIs) such as school/workplace closure, use of face masks, lockdown, quarantine, isolation, social distancing, and others became necessary to mitigate the pandemic [2]. A quantitative measure of these interventions to track the stringency or compliance of NPIs implemented by 139 country governments revealed an inverse relationship between the stringency index and case incidence in the first wave [2]. The COVID-19 pandemic exemplified how a population’s willingness to adopt infectious disease interventions is–increasingly–as important as accessibility, for supporting uptake of the intervention [3].

Mathematical models were used early in the pandemic to model the spread of severe acute respiratory syndrome coronavirus 2 (SARS-CoV-2), for instance to estimate *R*_0_ [4], assess the potential impacts of vaccination [5] and NPIs [6], and explore the role of spatial structure [7]. These analyses were able to offer a great deal without an endogenous representation of human behaviour in the models. However, being able to anticipate the public response to interventions–especially if adherence is partly or fully voluntary–could be useful for some application areas of models [8]. Thus, mathematically modelling the interplay between social dynamics and transmission dynamics (coupled behaviour-disease dynamics) during a pandemic cannot be overlooked [9, 10]. Multiple waves tend to emerge naturally from coupled behaviour-disease models [11]. Behaviour-mediated waves occurred not just for COVID-19 but also the’Spanish flu’ influenza pandemic of 1918-1919 [12], and thus are a possibility for future pandemics.

For both SARS-CoV-2 and other infectious diseases, modellers have explored how public awareness of the basic reproduction number (*R*_0_) played a significant role in reducing infection prevalence [13, 14]. However, the structure of groups within the population is also important: homophily (preferring to interact with those who are like you) and outgroup aversion (preferring not to interact with those who are not) can interact with disease characteristics like *R*_0_ to determine support for NPIs and thus the pattern of epidemic peaks [15]. Social learning (itself a type of ‘contagion’) is also highly influential [16]. Furthermore, access to accurate or inaccurate information, and the media, influence the dynamics of social behaviours that affect disease dynamics [17, 18, 19]. Coupled behaviour-disease models can show single or multiple peaks [20], and such models calibrated to empirical data performed comparably to non-behavioural models studied by the United States Centers for Disease Control [21]. Other COVID-19 models have evaluated pandemic interventions individually or in combination [20, 22], or studied the role of economic factors and information in decisions about whether to adopt an intervention [23].

Behaviour modelling can be either endogenous or implicit, and can take various approaches such as data-driven modelling, compartmental modelling of behavioural states, behaviour-informed transmission rate, or they can postulate separate state variables for behavior or opinions [20, 24]. Behavioural parameters can be inferred from time series or other data [23, 24] and the fitted model can then be used to forecast the disease dynamics [21, 24]. Comparative analysis of various coupled behaviour-disease models parameterised for different geographic locations has also been conducted [24]. Other approaches have compared forecasts of COVID-19 deaths in the United States for behavioural versus non-behavioural models [21].

To date, most of the literature on coupled behaviour-disease models is theoretical in nature, and relatively few analyses attempt to validate the models against empirical data or test the predictive powers of these models, including for COVID-19. Here we explore whether having case notification data from the first wave of the COVID-19 (Spring 2020) enables prediction of the second wave. We address the specific question: can a coupled behaviour-disease model predict the second wave better than a model that lacks a representation of population behaviour? To motivate our model design, we note that models do not need highly realistic structure in order to be useful. Simpler models may be suitable for exploratory analysis, conceptual development, or didactic purposes. More importantly, a highly realistic model can easily be over-fitted to available data, making it useless for prediction. To avoid overfitting, and because our goal was to assess whether adding endogenous behaviour to a transmsision model helps (not whether it is possible to achieve a good model fit, *per se*), we fit relatively simple models to case notification and pandemic stringency time series.

## Methods

We compared two models. One model is a standard Susceptible-Infectious-Recovered compartmental model with seasonality in the transmission rate, which captures how SARS-CoV-2 spreads through the population due to contacts (‘disease model’). The other model is based on social learning theory, according to which behaviour is learned from social influences [25]. The model adds a social learning mechanism to the disease model and allows a two-way interaction between SARS-CoV-2 transmission and social dynamics (‘coupled behavior-disease model’ or just ‘coupled model’). In the latter model, an increase in case incidence causes increased adoption of NPIs, which in turn causes a subsequent decline in cases to a degree depending on the intervention efficacy. Individuals are either mitigators or non-mitigators and they switch their behavior according to a utility function depending on the case incidence and cost of mitigation. Both models adjust for under-reporting of cases. Full details of both models are presented in the Supplementary Appendix (pp 1 – 8).

We assessed the predictive powers of our coupled behaviour-disease model against the disease model, by fitting the models to the first COVID-19 wave of Spring 2020 in thirteen European countries (see Supplementary Appendix pp 5 – 6 for inclusion/exclusion criteria), and seeing how well the models predicted the second COVID-19 wave in those countries. For model parameter inference, we used an approximate Bayesian computing sequential Monte Carlo method. This method yields a ‘cloud’ of inferred parameter sets (particles) that all achieve a similarly close agreement with the data. These posterior distributions can be used for subsequent analysis, such as (in our case) generating predictions for the second COVID-19 wave. Because we are only using time series of case notifications and a stringency index representing the aggregate effect of multiple pandemic interventions, we keep both the disease model and the coupled model as simple as possible, to prevent over-fitting the model. The disease model was only fitted to COVID-19 case notification data, while the coupled model was fitted to both case notification and stringency index data. The data used in the study are shown in Figure 1, and full details of the analysis appear in the Supplementary Appendix pp 4 – 9.

**Figure 1.**
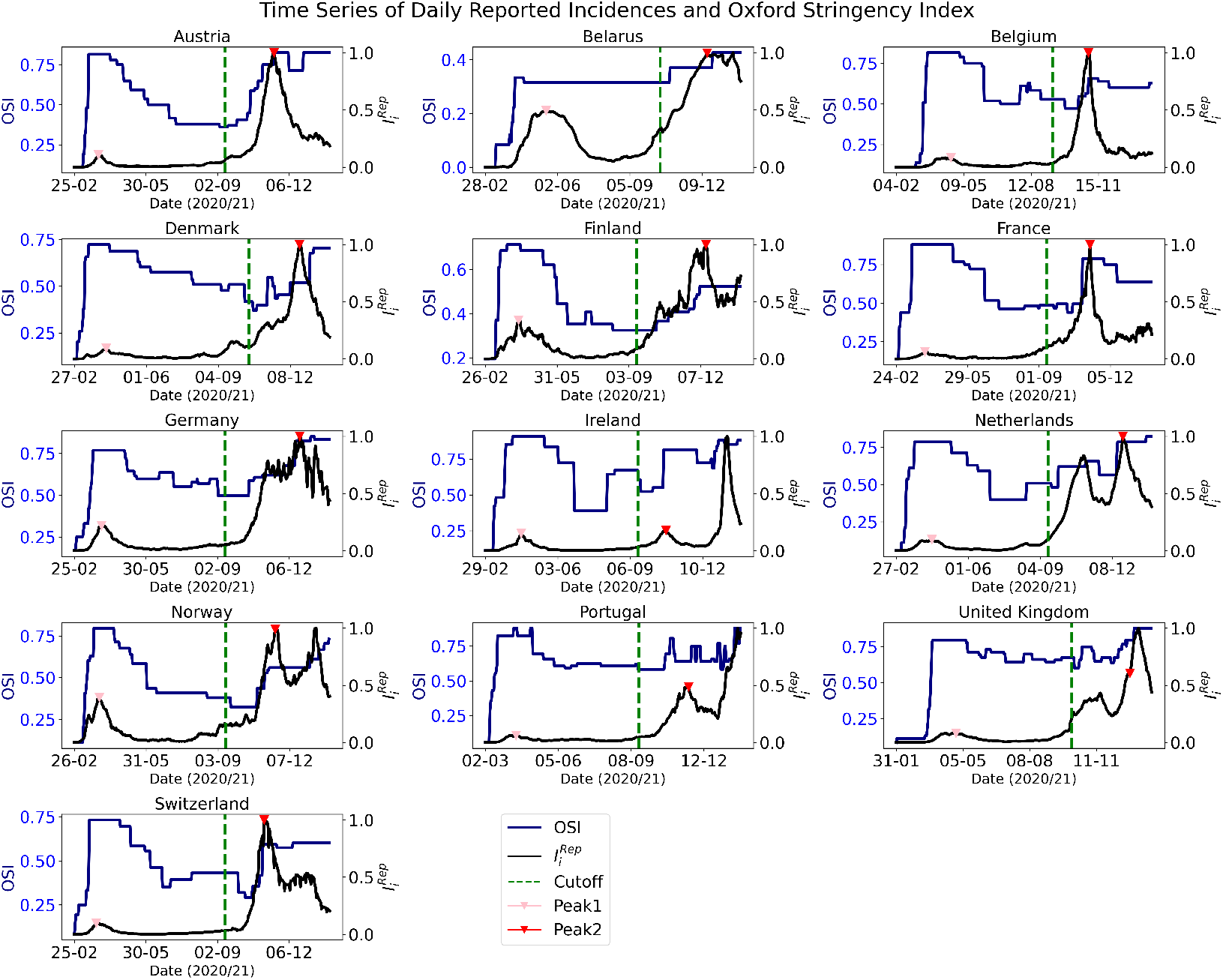
Empirical data used in study. Panels show daily reported cases 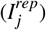 and corresponding stringency index (OSI) for 13 countries in Europe. The green line splits the data into two parts: first wave, used for fitting, and second wave, which is for retrospective prediction. The first peak (pink triangle) is defined as the maximum number of cases reported between March and May. The second peak (red triangle) is the maximum number of cases in second wave from September to December.

## Findings

### Model fits to the first wave

The posterior distribution of the inferred parameters falls within acceptable thresholds for both models (see Supplementary Appendix pp 5). Violin plots of the posterior parameter sets for both models show the expected clustering around median values, with some tail-end dispersion (see Supplementary Appendix pp 18 – 22). However, the particles (parameter sets) for the disease model have a noticeably lower mean squared error than the coupled model, because they have fewer free parameters (Supplementary Appendix, Table S2). The disease model fits the data better than the coupled model for Belarus, Belgium, Denmark, Finland, France, Germany, the Netherlands, Portugal and the United Kingdom, according to three statistical metrics (Supplementary Appendix Table S4). Our results from model fitting (‘training’) confirm both the disease model and the coupled model as adequate fits, with clearly identifiable parameter ranges, but the fit for the disease model is somewhat better than the coupled model.

### Model predictions of the second wave

As a result of the better fit of the disease model during the first wave, one might expect it to predict the second wave better than the coupled behaviour-disease model. However, it actually does worse, because it does not account for interactions between population behaviour and disease dynamics (Figure 2). When cases fell after the first wave, populations relaxed control measures. But eventually, this created the conditions for a second wave. In the face of rising case notifications in this second wave, populations changed their behaviour and increased their adoption of non-pharmaceutical interventions, which slowed down viral transmission. The coupled model tracks this ebb and flow of behaviour and case incidence through the utility function, which expresses population preferences based on current case notifications. Since the disease model does not include an endogenous response that could slow down the spread of infection, it predicts a sharp and dramatic increase in case incidence during the second wave that exceeds what actually happened in most countries (Figure 2, black line). Whereas, the coupled model predicts a lower peak that is better aligned with the empirical case notifications, on account of including the feedback from behavior (Figure 2, orange line). However, the 95% credible intervals for the coupled model are quite broad, which is to be expected given the inherent uncertainties in the temporal evolution of a coupled behavior-disease system.

**Figure 2.**
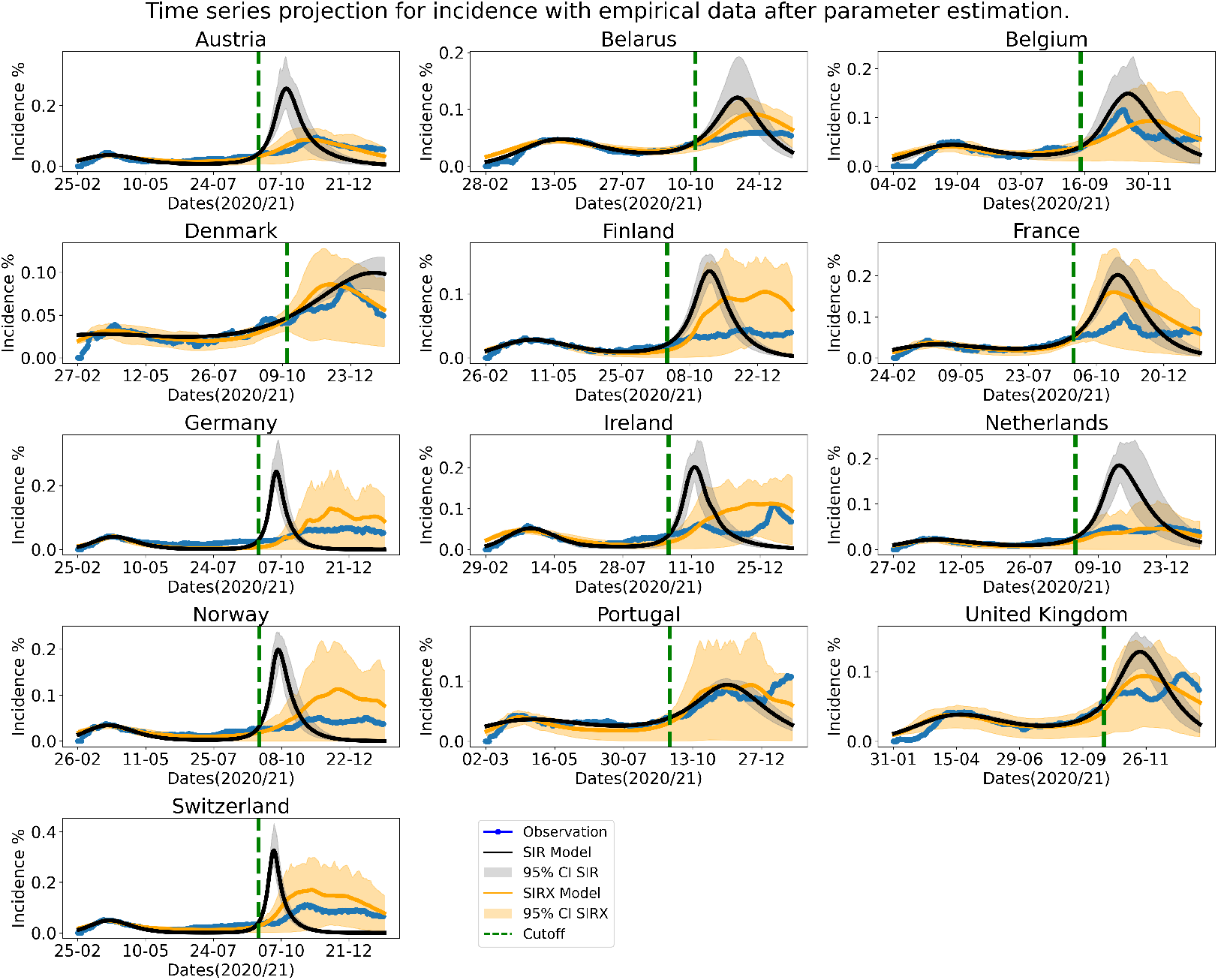
Predictions of the coupled behaviour-disease model for the second wave are closer to the empirical data. Trajectory of the disease model (SIR) and the coupled behaviour-disease model (SIRX) from estimated particles from the model(s) training. We show the actual observation, average simulation and the 95% credible interval. The green vertical dashed-line divides the time series into the training (fitting) portion during the first wave, and the predicted portion for the second wave.

We also compared the model prediction of the peak magnitude (defined as the highest daily case notifications during the second wave) and peak day (the day on which the peak occurred). The median peak magnitude across the 13 countries for the coupled model is much closer to the empirical peak magnitude, than is the case for the disease model (Figure 3a), reflecting the slowing effects of behavioral feedback, as already discussed. However, neither model is universally effective in predicting the peak day. The coupled model predicts the peak day within a margin of error of 7 days only for Austria, Belarus and the Netherlands, while the disease model is accurate within the same margin for Belgium, Ireland and Portugal (Figure 3b). The peak day in the empirical data usually falls outside the 95 % credible interval for the disease model. The opposite is the case for the coupled model, although the credible intervals for the coupled model are too broad to expect an accurate prediction.

**Figure 3.**
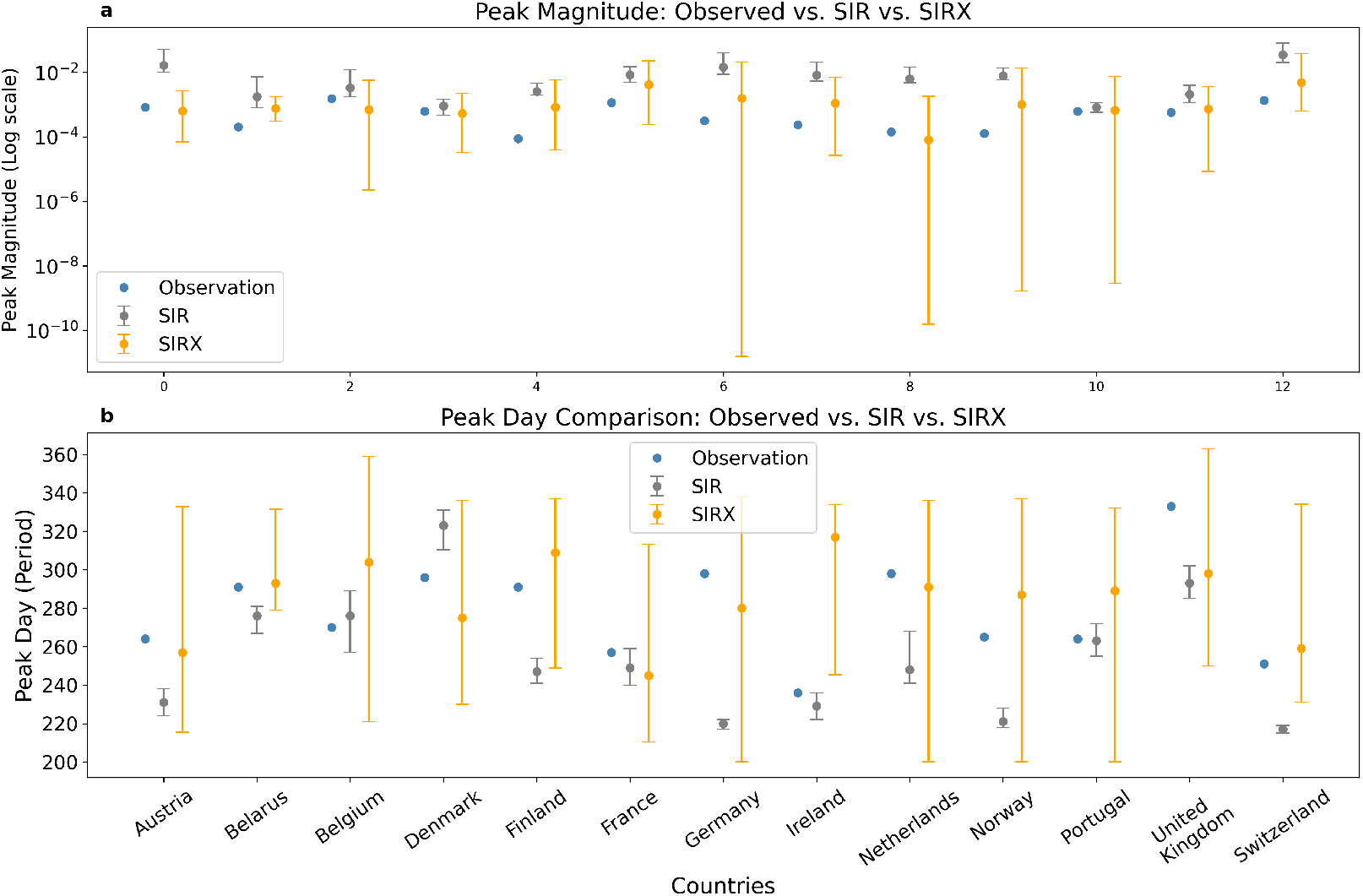
Predicted peak magnitude for the coupled behaviour-disease model is closer to the empirical data, though not the predicted day of the peak. Actual peak magnitude (**a**) and peak day (**b**) compared to model estimates. We show the identified peak details and compare with estimation from disease model (SIR) and coupled behaviour-disease model (SIRX) with 95% credible interval.

### Behaviour Dynamics for Coupled Model

The coupled model generates not only case notifications, but also a behavioural state variable–the proportion of the population that adopts non-pharmaceutical interventions. Over the first wave, the coupled model was fitted to both case notifications and the Oxford stringency index, which measures how many different interventions are being applied in each country. Our implicit assumption is that it is possible to equate our data source–the Oxford stringency index–with our model output variable–the average uptake of NPIs in the population. While they will not be perfectly correlated, we note that these two measures were largely consistent in the first two waves of the pandemic, and especially in the ‘high-trust’ societies that characterise most of the countries we studied [26, 27].

The median fit of the coupled model tracks the stringency index with good qualitative accuracy for 11 out of 13 countries, in terms of capturing the initial surge in mitigation, followed by the relaxation of measures later in the first wave, and the subsequent re-application of measures in the second wave (Figure 4). However, the coupled model could not be fitted to Belarus or Finland. The predicted case incidence was also relatively high for those two countries, as might be expected (Figure 2). Since the two countries did not fit the Oxford Stringency Index, the coupled model effectively does not account for behaviour (the proportion of predicted mitigators is zero). This resulted in an increase in case incidence similar to the disease model compared to the actual. The 95% credible interval contains the empirical data except for Belarus and Finland. The 95% credible interval is more broad for the behavioural data on account of the lower weighting used for behavioral data when fitting the coupled model to both case notification and stringency index datasets. Despite the broad credible intervals and failure for Belarus and Finland, the model’s qualitative prediction of the revival of NPIs during the second wave is correct. In each country except for Belarus, Finland and Ireland, the predicted population support for NPIs increases during the second wave, following the same trend as the stringency index (Figure 4).

**Figure 4.**
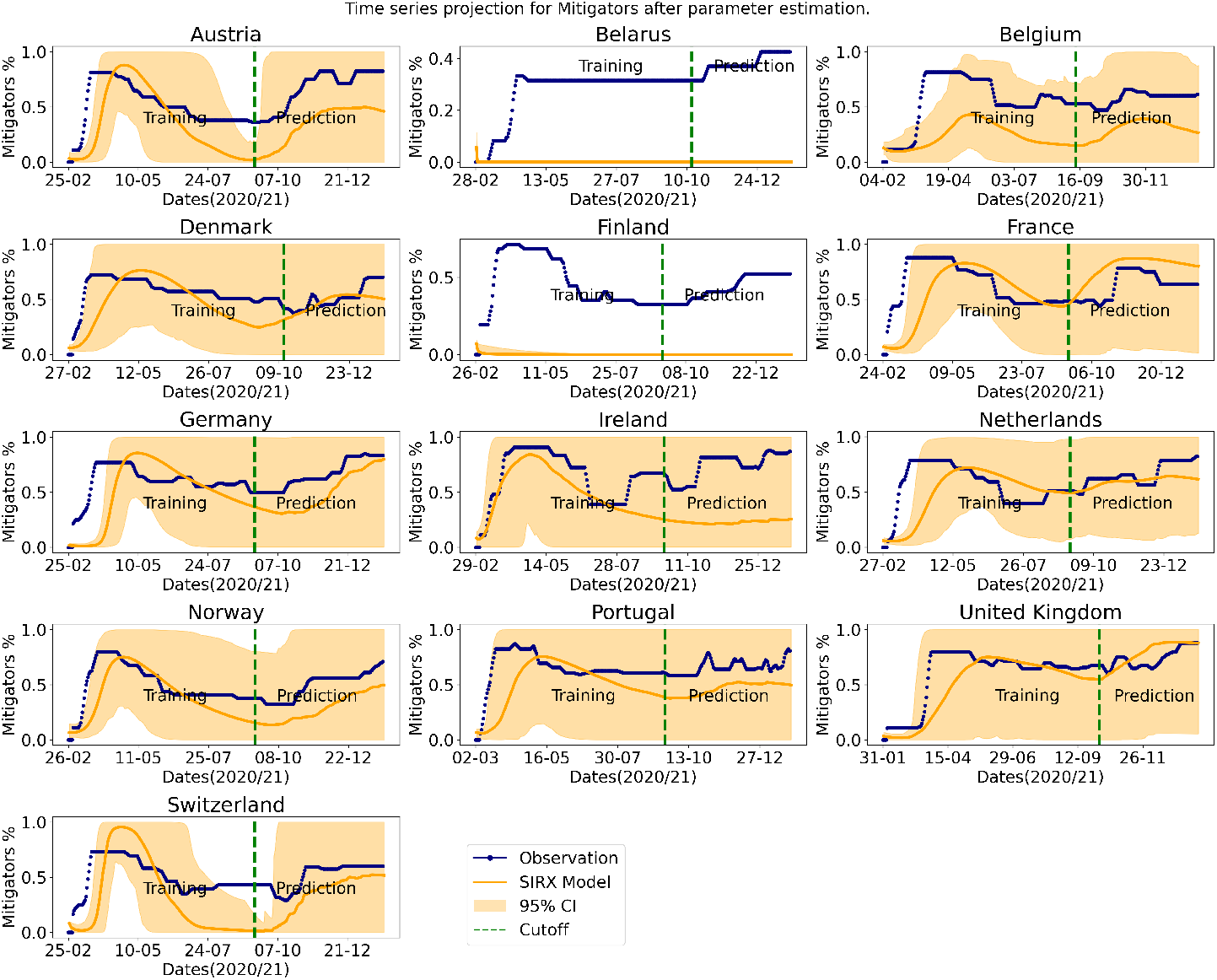
Coupled behaviour-disease model predicts qualitative trends in adoption of NPIs. Trajectory of the behavioural prediction of the coupled behaviour-disease model (SIRX) fitted to the Oxford Stringency Index simultaneously with daily reported cases. We show the actual observation, average simulation and the 95% credible interval.

### Statistical analysis

The area between the plotted empirical and modelled case notifications was computed for both models. A smaller difference in area means the model remained closer to the empirical data over the course of the pandemic wave. Across all 13 countries, the coupled model has a smaller area than the disease model, implying that the predicted curve of the coupled model remained closer to the empirical data, than the disease model (Figure 5a).

**Figure 5.**
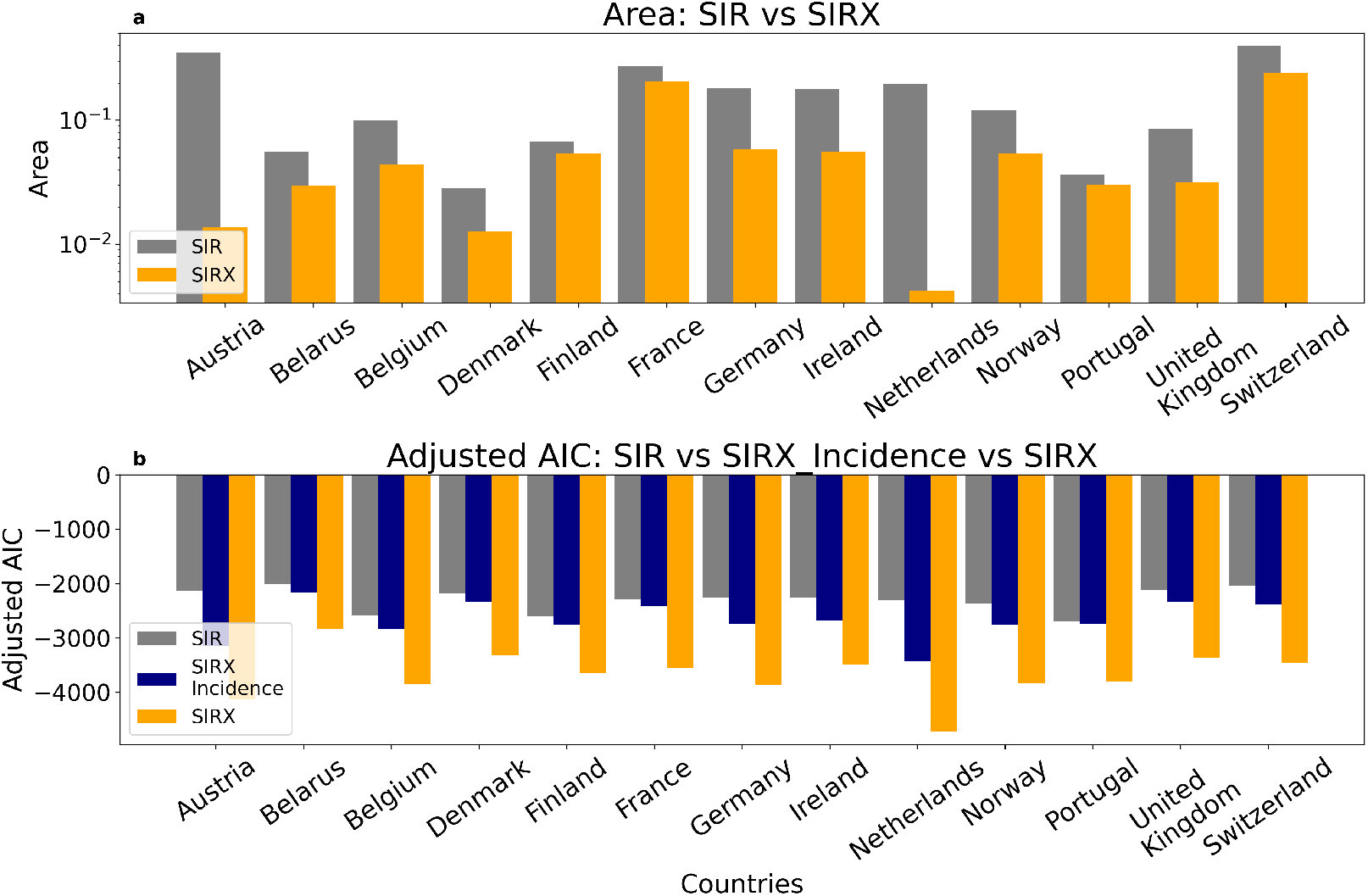
Coupled behaviour-disease model explains the data better, according to statistical metrics. (**a**) Adjusted AIC for disease (SIR) and coupled behaviour-disease (SIRX) – incidence (blue), and incidence + stringency (orange), models for the prediction period of the empirical data. (**b**) Area between the untrained data and our model predictions for the incidences.

Our findings were further evaluated using the adjusted Akaike Information Criteria (AICc). This information theory metric selects which of several models best explains the data, defined as a high goodness-of-fit without requiring too many fitted parameters. For the disease model, we calculated this metric using the goodness-of-fit against case notification data. For the coupled model, we calculated it using goodness-of-fit for both the case notification on its own, and the case notification plus the stringency index data. The coupled model was penalized with the full number of fitted parameters (i.e., both behavioural and epidemiological parameters), even when the goodness-of-fit was computed only using case notification data. This approach provided a more conservative test of the coupled model. Portugal recorded the lowest difference for the adjusted AIC; we observe same country has the lowest difference for the area whilst Austria records the highest difference for the area (Figure 5).

The analysis shows that the AIC for the disease model is higher (more positive) than the AIC for the coupled model, regardless of which way the AIC is computed for the coupled model, and for all 13 countries (Figure 5b). A higher score indicates that the model explains the data less well. The strongest difference between the model is for the Netherlands where the AIC for the coupled model is significantly lower than for the disease model, as the predicted case notifications are significantly higher than the empirical data.

These two statistical analyses support the hypothesis that the coupled model explains the data better than the disease model. The AIC results are especially notable, given that the behavioural model has a significantly larger number of parameters than the disease model, and was penalised more strictly.

The basic (*R*_0_) and effective(*R*_*e f f*_) reproduction numbers were also estimated (Supplementary Appendix Table S3). *R*_0_ was estimated for the coupled model with mitigation ‘turned off’, and for the disease model. *R*_*e f f*_ was computed from the coupled model with mitigation ‘turned on’. *R*_*e f f*_ was lower than *R*_0_ for the disease model, reflecting the impact of mitigation on SARS-CoV-2 transmission.

### Interpretation

This study achieves two important key outcomes: it supports the role of feedback between population behaviour and disease dynamics, and it shows that including this feedback loop in transmission models can improve their ability to predict and explain observed case incidence data, in our COVID-19 study system. Without a population behavioural response to slow the spread of the virus, the standard compartmental model predicted an unrealistically large and quick second pandemic wave.

In other words, these results support a perspective where SARS-CoV-2 dynamics and population behaviour should not be viewed in isolation from one another, but rather as two parts of a single, coupled behaviour-disease system. This view is in line with a school of thought that human populations are complex, adaptive, and self-organized systems consisting of interacting agents [28]. In such cases, public health authorities must contend not only with how effective and safe an intervention is, but also how the population will respond. For instance, the term ‘policy resistance’ has been coined to describe situations where the population response to a public health intervention serves to partially undermine the intervention [29].

In more concrete terms, our results show how coupled behaviour-disease models can add to the public health toolbox for the control of infectious diseases. Standard transmission models that do not account for behaviour will always be useful, such as for estimating the basic reproduction number *R*_0_ early during a pandemic. However, coupled behaviour-disease models can serve complementary roles. For instance, public health authorities could use such models anticipate a population’s reaction to various possible infection control measures, and how the population uptake of such measures will vary with case incidence. In this way, public health authorities could attempt to identify risk factors for public reactions that undermine proposed public health measures, and implement measures that are most likely to be effective in reducing morbidity and mortality, given what is known about the behavioural landscape of the population.

For this to happen, coupled behaviour-disease models with greater structure must be validated using a growing suite of traditional and digital datasets on opinion and behaviour [9]. For this study, we intentionally used simplistic models, in order to avoid the risk of over-fitting and thus losing predictive power. Some of the simplifying assumptions that we made could be relaxed in future work. For instance, the model could be stratified with respect to the different types of possible interventions–each with its own specific mitigation cost–so that support for specific measures could be studied. And, the perspectives and costs specific to public health authorities (sanitation cost, subsidies, loss of tax revenue) could be included to improve the usefulness of coupled models for decision support. Similarly, other sources of behavioural data could be used in combination, such as mobility data from GPS tracking, and sentiment data from online social media or Internet searches [30]. In summary, our results show that adding behavioural modelling to infectious disease transmission models can improve their ability to explain and predict the early waves of the COVID-19 pandemic. The improvement demonstrates the potential of coupled behaviour-disease models not only for explaining epidemiological patterns in populations, but also as a potential tool for designing public health interventions during a pandemic.

## Data Availability

All data produced are available online at https://github.com/SefahF/Pandemic-waves-as-the-outcome-of-coupled-social-and-disease-dynamics

## Data sharing

The code and data extracted for this study are publicly available and can be accessed here source code. The main data source was obtained from [OurWorldindata](https://ourworldindata.org/coronavirus).

## Contributors

CTB conceived the model and SF established the results. SF drafted the manuscript and CTB reviewed and edited the manuscript. Both authors read and approved the final version of the manuscript. SF accessed and verified the data used in the study, and had final responsibility for the decision to submit for publication.

