## Supplementary Appendix for "Pandemic waves as the outcome of coupled behaviour and disease dynamics: a mathematical modelling study"

### Supplementary Appendix: Pandemic waves as the outcome of coupled social and disease dynamics: a mathematical modelling study

Sefah Frimpong, Chris T. Bauch

*University of Waterloo, Department of Applied Mathematics, Waterloo, N2L 3G1, Canada*

#### Supplementary Methods

**Model Overview.** Many infectious diseases like SARS-CoV-2 largely transmit from an infected person to a healthy person and have several consequences with an extreme case of death. We consider a compartmental model where the population is categorised into Susceptible, Infected, and Recovered groups – susceptible group is the healthy population but have high probability of getting infected with the virus, the infected group is the proportion of the population who have been infected by the virus and have also become infectious(agent of transmission), and recovered group is the proportion of the population who have either recovered from the virus or died as a result of infection. The population takes preventive measures such as the use of NPIs to protect themselves and others which as a consequence have significant effect on the susceptible and infected group. However, due to the longevity of COVID-19 virus existence, some of the population readjust their strategies per unit time.

**Disease Transmission Model.** We formulate a seasonal SIR (susceptible, infected and recovered) compartmental model. We consider a dense population with many susceptible to the disease as there was very low/little immunity against the COVID-19 virus in the early stages of the disease outbreak. Susceptible individuals who come into contact with the virus or infected surfaces become infected and infectious in a very short period (1-7 days on average) [20]. Infected individuals either die or recover thus moving to the recovered compartment. Susceptibility and infections are affected by seasonal variations of weather conditions such as winter, spring, summer, and fall due to 1) a change in the survivability of the virus and 2) changes to contact patterns of individuals within the year [2, 6]. The compartments are expressed using a conventional approach of differential equations to represent the rate of change in each group with respect to time(day) while the seasonal variation is defined using a sinusoidal function [6]. The transmission dynamics given by an SIR model with seasonality is presented below;

$$\frac{dS}{dt} = -\beta(1 + b\cos(t-\phi))SI, \tag{1}$$

$$\frac{dI}{dt} = \beta(1 + b\cos(t-\phi))SI - \gamma I, \tag{2}$$

$$\frac{dR}{dt} = \gamma I, \tag{3}$$

where $S$ is the proportion of susceptible individuals ("susceptible"), $I$ is the proportion of individuals who are both infected and infectious ("infectious"), and $R$ is the proportion of individuals who are no longer infectious ("removed"). $\beta$ is the baseline transmission rate in the absence of preventive strategies (NPIs), $b$ is the amplitude of seasonal transmission, $\phi$ is the phase of seasonality, and $\gamma$ is the time rate at which an infectious person recovers or is removed from the population.

**Behaviour Model.** The behaviour of individuals during epidemics (pandemics) contribute to the outcome of the prevalence of the pathogen and morbidity of infectious. Individual responses to SARS CoV-2 (COVID-19 virus) have not been any different to the previous disease outbreaks. Persons either make changes to their behavioural patterns (that is, their daily routine changes) to keep themselves and their family safe or risk their health to live as no disease outbreak had occurred. In the absence of effective and efficient vaccines to control the virus, Non-pharmaceutical interventions (NPIs) becomes the appropriate public health strategy to adopt in controlling SARS CoV-2 virus. This strategy has been widely implemented in most countries and have been significant in its objective [12]. We build a behavioural dynamic model base on individual strategy to adhere or not adhere to the use of NPIs as the individual aims for the best available strategy (that is, the strategy with the highest payoff) and couple it with our conventional SIR model in a similar approach as [3, 17].

In building the behaviour model, we use the concept of evolutionary game theory involving imitation dynamics by considering two players corresponding to the two strategies; support for NPIs (Mitigators) strategy and no support for NPIs (Non-mitigators) strategy. We assume that all information is available to every player and the perceived payoff for the players in each group are the same,that is, mitigators have the same perceived payoff and non-mitigators have the same perceived payoff. Let $x$ represent the proportion of Mitigators and $(1-x)$ as the proportion of Non-mitigators. For each strategy, there is a perceived reward associated with it. We consider rather simple yet realistic payoffs. The perceived payoff for Mitigators ($E_s$) is;

$$E_s = -c \tag{4}$$

where $c$ is the cost of maintaining NPIs measures (such as the provision of relief funds for persons out of work, use of face mask, etc). We assume that there is no direct threat to mitigators, such as morbidity from NPIs, except for the cost associated with its implementation. Similarly, the perceived payoff for Non-mitigators ($E_n$) is;

$$E_n(I) = -I\exp(-\lambda t) \tag{5}$$

where $\lambda$ quantifies the sensitivity to changes in prevalence (how the population learns from the virus and treatment development), $\exp(-\lambda t)$ is the perceived probability of morbidity from not adhering to NPIs. This is also expressed as a function of the prevalence which implies that a low level of cases corresponds to low payoff and high level of cases will lead to high payoff. We note here that there are no gains (income or benefits) to any of the players except cost (monetary or morbidity from infection). The lower the payoff, the better for the players.

As the disease progresses, individuals switch strategies to adopt the best by sampling the population and imitating their decisions. An imitation is successful when one strategy is substituted for the other based on the difference in perceived payoff. For instance, when $E_s - E_n(I) > 0$, then support for NPIs is a preferred strategy which implies non-mitigators imitate mitigators while $E_s - E_n(I) < 0$ implies support against NPIs is the strategy preferred meaning mitigators become non-mitigators. The rate of switching from non-mitigator to mitigators strategy is formulated using the updating scheme of proportional imitation rule;

$$\frac{dx}{dt} = kx(1-x)\rho(E_s - E_n(I)) \tag{6}$$

where $k$ is the time rate at which individual samples are made from a population and $\rho$ is the constant of proportionality since $\rho(E_s - E_n)$ is the probability of non-mitigators switching to mitigators.

Similarly, when $E_s - E_n(I) < 0$ then the rate of switching from mitigator to non-mitigator strategy is;

$$\frac{dx}{dt} = -kx(1-x)\rho(E_n(I) - E_s) \tag{7}$$

We simplify the expression by taking $\kappa = k\rho$, thus (equation 6) then becomes;

$$\frac{dx}{dt} = \kappa x(1-x)(I\exp(-\lambda t) - c) \tag{8}$$

where we consider the quadratic term $x(1-x)$ to represent a social learning dynamics which individuals sample others at some probability rate, and they change opinion based on the payoff difference $(E_s - E_n(I))$ [17]. This behaviour dynamic model (equation 6) considers the social impact of COVID-19, the rate of social learning, and the cost involved in ensuring that the population adheres to the interventions implemented.

**Coupled Behaviour-Disease Model.** The rate of support, that is mitigators ($x$) for the use of NPIs affects the proportion of susceptible ($S$) and infectious ($I$) groups in two ways; 1) individuals protect themselves by playing mitigator strategy at the very beginning thus influence the number of susceptible 2) individuals begin with a non-mitigator strategy gets infected and switches to mitigator strategy to protect their families and loved ones (eg, agreeing to isolate and quarantine) which then influences the prevalence of the disease. As much as NPIs play a significant role in disease outbreak, the efficacy of their application is equally important. For instance, hand washing must be effectively done to achieve its desired result. We introduce a parameter $\epsilon$ to represent the efficacy of the use of NPIs which is expressed to be proportional to the rate of support $(\epsilon x)$ giving us a better representation of the effect of mitigator strategy on the disease dynamics. The complement $(1-x\epsilon)$ which now represent the proportion of non-mitigator strategy make it to the susceptible and infectious compartment. The coupled model is then constructed by combining the disease model and behaviour model. We multiply the proportion of susceptible and infected rates with respect to time(day) of the conventional disease model with $(1-x\epsilon)$, representing social learning dynamics. This results in the following model equations for the coupled disease-behaviour model.

$$\frac{dS}{dt} = -\beta(1+b\cos(t-\phi))(1-x\epsilon)SI, \quad (9)$$

$$\frac{dI}{dt} = \beta(1+b\cos(t-\phi))(1-x\epsilon)SI-\gamma I, \quad (10)$$

$$\frac{dR}{dt} = \gamma I, \quad (11)$$

$$\frac{dx}{dt} = \kappa x(1-x)(I\exp(-\lambda t)-c) \quad (12)$$

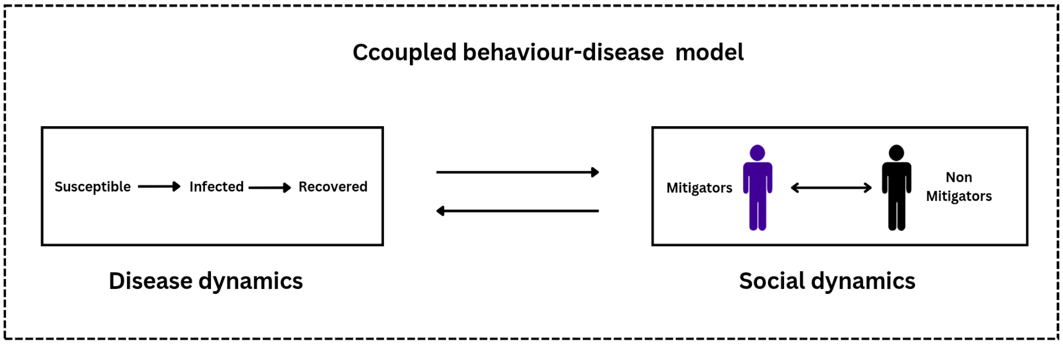


**Fig. S 1: Schematic diagram for model.** Feedback loop between disease dynamics and social dynamics for Coupled behaviour-disease.

**Daily Incidence.** We estimate the daily incidence from the model(s) by first estimating the cumulation of cases at time $(t)$ and previously confirmed cases. Daily incidence is defined as the daily confirmed cases at a specific time $(t)$, that is, the number of individuals estimated to have been infected with the virus at a particular time $(t)$. We let $C$ represent the proportion of cumulative cases, thus, the daily incidence $(I_i)$ is defined as the difference between cumulative infections $(C)$ from one day $(t)$ to the next $(t+1)$. Since we have two models, we have different expressions for $C$ and for representation purposes $C_{\text{SIR}}$ and $C_{\text{SIRX}}$ are estimated cumulative cases for SIR (disease model) and SIRX (coupled model), respectively. The proportion of cumulative cases $(C_{\text{SIR}})$ and $(C_{\text{SIRX}})$ modeled as a time derivative are presented below;

$$\frac{dC_{\text{SIR}}}{dt} = \beta(1+b\cos(t-\phi))SI \quad (13)$$

$$\frac{dC_{\text{SIRX}}}{dt} = \beta(1+b\cos(t-\phi))(1-x\epsilon)SI \quad (14)$$

With the knowledge of under-reporting of cases, we introduce a parameter ($\eta(t) > 1$) [14] which accounts for low reporting of confirmed cases. This is then factored into the estimation of daily incidences from the model(s). We formulate the daily incidence at time ($t$) as;

$$I_i(t) = \eta(t)^{-1}(C(t) - C(t-1)) \quad \text{where } I_i(t) = \{Ii_{\text{SIR}}(t), Ii_{\text{SIRX}}(t)\} \text{ and } C(t) = \{C_{\text{SIR}}(t), C_{\text{SIRX}}(t)\} \quad (15)$$

This is then fitted to the case notifications – daily incidence reported ($I_i^{\text{rep}}(t)$) from the COVID-19 observation case notifications to obtain the posterior distribution of the model parameters. We discuss next parameter estimation (fitting of the model to empirical data).

**Parameterisation.** Parameter estimation is performed using an approximated Bayesian statistics where the posterior distribution is estimated using the prior distribution (here as assume uniform distribution) and the likelihood with respect to the empirical data since we do not have information on the marginal probability. This is illustrated below;

$$\text{posterior} \quad \propto \quad \text{prior} \times \text{likelihood} \quad \{\text{over the empirical data}\} \quad (16)$$

$$\pi\left(\theta^{**}\right) \quad \propto \quad \pi\left(\theta\right) \times \pi\left(\theta | x_0, y_0\right) \quad (17)$$

where $\theta$ represent parameter, $\theta^{**}$ represent estimated parameter, $\pi(\theta)$ represent prior distribution (uniform distribution), $\pi(\theta|x_0, y_0)$ represent likelihood over the data $x_0$ and $y_0$, and $\pi(\theta^{**})$ represent posterior distribution of estimated parameter.

This concept forms the foundation of our parameter estimation technique. To implement this, we use Approximated Bayesian Computation Sequential Monte Carlo (ABC-SMC). This is used for both parameter estimation and model selection. The method uses the concept discussed earlier, but in a sequential manner, to obtain meaningful and reasonable parameter values that are identifiable with the empirical data. The algorithm for ABC-SMC is discussed by [19] which is implemented in this study.

To implement ABC-SMC, we sample the parameter values from their prior (uniform) distribution obtained through inference, then apply the estimated parameter values to the model equation to obtain its simulated daily incidence ($I_i$). The distance between the simulations($I_i$) and data ($I_i^{\text{rep}}(t)$) is evaluated and assessed using the tolerance level. We accept the estimated parameter values when the distance is less than or equal to the tolerance and reject otherwise with a corresponding weight for the particles. This is repeated for a number of steps (iterations) and a corresponding adjustment to the tolerance level to obtain a good fit (parameter estimates). We estimate at least 1000 particles (parameter set) per iteration for at most 31 iterations. This gives us particles from at least $15,000$ to $31,000$ that make up the posterior distribution for each parameter. We are unable to run more particles for the coupled model due to its complex nature. Also we had two tolerance levels sets 1) disease tolerance 2) behaviour tolerance. However, for the disease model we are able to run more particles due to its less complex nature and with one tolerance level set.

**ABC SMC algorithm.** For parameter estimation, we use the Approximated Bayesian Computation Sequential Monte Carlo algorithm discussed by [19] with a few modification for our use. The algorithm is presented below;

| | |
|---|---|
| Step 1 | Start $\epsilon_{I_1}, \ldots, \epsilon_{I_T}$. |
| | $\epsilon_{x_1}, \ldots, \epsilon_{x_T}$. |
| | Set the population indicator $t = 0$. |
| Step 2 | Set the particle indicator $i = 1$. |
| Step 2.1 | If $t = 0$, sample $\theta^{**}$ independently from $\pi(\theta)$. |
| | Else, sample $\theta^*$ from the previous population $\left\{\theta_{t-1}^{(i)}\right\}$ with weights $w_{t-1}$ and perturb the particle to obtain $\theta^{**} \sim K_t(\theta \mid \theta^*)$, where $K_t$ is a perturbation kernel. |
| | If $\pi(\theta^{**}) = 0$, return to Step 2.1. |
| | Simulate a candidate dataset $I^*, x^* \sim f(I, x \mid \theta^{**})$. |
| | If $d(I^*, I_0) \geq \epsilon_{I_t}$ or $d(x^*, x_0) \geq \epsilon_{x_t}$ , return to Step 2.1. |
| Step 2.2 | Set $\theta_t^{(i)} = \theta^{**}$ and calculate the weight for particle $\theta_t^{(i)}$ |
| | $w_t^{(i)} = \begin{cases} 1, & \text{if } t = 0 \\ \frac{\pi\left(\theta_t^{(i)}\right)}{\sum_{j=1}^{N} w_{t-1}^{(j)} K_t\left(\theta_{t-1}^{(j)}, \theta_t^{(i)}\right)}, & \text{if } t > 0 \end{cases}$ |
| | If $i < N$, set $i = i + 1$, go to Step 2.1. |
| Step 3 | Normalize the weights. If $t < T$, set $t = t + 1$, go to Step 2.0. |

**Implementation of Algorithm.** The algorithm is implemented as presented above with focus on the choice of the threshold for each population, sampling from the prior distribution and the number of iterations ($T$). The threshold is initialsed then updated for each iteration (simulation) for the incidence and behaviour separately;

(A) Incidence: Initialise $\epsilon_{I_0}$ then $\epsilon_{I_t} = \max\{\varepsilon_1 \ldots \varepsilon_N\}$, where $\varepsilon_i$ is the mean squared error $(d(I^*, I_0))$ for each estimated particle $(\theta^{**})$ for $i = 1, \ldots, N$ set of particles ($N = 1000$). This is run for $T$ times giving us a total set of particles of $N \times T$.

(B) Behaviour: Initialise $\epsilon_{x_0}$ then $\epsilon_{x_t} = ||\epsilon_{x_{t-1}} - 0.005||$ for the population indicator $t = 1, ..., T$.

We check these threshold for every set of particles simultaneously to select acceptable particles. For SIRX model, we set $T = 15$ and for SIR model $T = 25$. This was done because of the difference in complexity of the two models as SIRX model was more expensive than the SIR model. These $T$ values gave us very good results for the respective models.

- **S Code 1:** The code and data extracted for this study can be accessed here source code.

**Data.** To both estimate and validate our models, we obtain empirical data already collected on COVID-19 case notifications and corresponding changes in population behaviour. For the disease dynamics, we use the daily confirmed (reported) cases and for behaviour we considered the Oxford Stringency Index. The daily confirmed cases enables us to know the transmission (projection) of the virus in the population which we match with our daily incidence in the model. The Oxford Stringency Index gives the idea of government support for the use of NPIs and level of implementation. This influences the behaviour of the population as a whole and not necessarily at the individual level. Although public health policies of NPIs backed by governments can be said to account for such change in individual behaviour is reflected in the Google mobility data. Generally these give us the information we need to understand the disease dynamics and population behaviour change and achieve the motive for this study. We focus on the first year of the SARS-CoV-2 transmission in 2020 and early 2021.

Since every country in the world reported cases of the COVID-19 virus, we needed to narrow down on some of the countries based a number of factors which are listed below for the purpose of the study. These factors form the selection criteria for our case study.

- We centered are study on countries in Europe due to availability of data and better quality compared with other countries.

- These countries must be performing frequent daily tests at least 100 tests per a 1000 of the population as of April 6, 2021 date of collection of the data, that is, total tests/1000 $\geq$ 100.

- The population size of the countries must be at least 3 million.
- The trajectory of COVID-19 reported cases for the country must have between 1 to 3 major peaks.
- The major first peak should have occurred during Spring (March to May) and second or third major peak should have occurred during Fall (September to November/December).
- The use of non-pharmaceutical interventions such as closure of schools and work places, wearing of mask, hand washing, social distancing among others should have been implemented at the latest middle of Spring 2020.
- **S Data File 1:** The main data source was obtained from OurWorldindata.

The prediction period for the second wave was chosen after the trough between first and second waves, and after the case incidence began to increase again (September/October depending on the country).

The table below gives a summary of the countries selected and length of data for our case study;

**Table S1. Length of data points for each country with start, cutoff and end dates.**

| **Country** | **Start Date** | **End Date** | **Cutoff Date(Length)** | Data Length |
|---|---|---|---|---|
| Austria | 25-02-20 | 28-01-21 | 12-09-20 (200) | 339 |
| Belarus | 28-02-20 | 28-01-21 | 15-10-20 (230) | 336 |
| Belgium | 04-02-20 | 28-01-21 | 11-09-20 (220) | 360 |
| Denmark | 27-02-20 | 28-01-21 | 14-10-20 (230) | 337 |
| Finland | 26-02-20 | 28-01-21 | 13-09-20 (200) | 338 |
| France | 24-02-20 | 28-01-21 | 11-09-20 (200) | 340 |
| Germany | 25-02-20 | 28-01-21 | 12-09-20 (200) | 339 |
| Ireland | 29-02-20 | 28-01-21 | 16-09-20 (200) | 335 |
| Netherlands | 27-02-20 | 28-01-21 | 14-09-20 (200) | 337 |
| Norway | 26-02-20 | 28-01-21 | 13-09-20 (200) | 338 |
| Portugal | 02-03-20 | 28-01-21 | 18-09-20 (200) | 333 |
| United Kingdom | 31-01-20 | 28-01-21 | 07-10-20 (250) | 364 |
| Switzerland | 25-02-20 | 28-01-21 | 12-09-20 (200) | 339 |

**Table S** 1: Summary of start, cutoff (data points fitted) and end date extracted for 13 European countries selected for case studies.

**Parameter and Initial Values Range.** Parameter estimation is carried out by assuming ranges for the prior distribution from which sampling is made. Parameter values including the initial values for the state variables are estimated, and the table below summarises all the parameters and initial values with their respective ranges.

**Table S2. Parameter names, range and their sources**

| Parameter | Meaning | Range | Source |
|---|---|---|---|
| $\beta$ | Transmission rate | 0.1 - 0.9 | $\beta = R_0 \times \gamma$ [8] |
| $b$ | Seasonal transmission amplitude | 0 - 1 | [9] |
| $\phi$ | phase of seasonal transmission | $-90°$ - $90°$ | [9] |
| $\epsilon$ | Intervention effectiveness | 0.1 - 0.8 | [13] |
| $1/\gamma$ | Infectious period | 2 - 10 days | [4, 16] |
| $\kappa$ | Social learning rate | 100 - 10,000/day | [5, 14] |
| $\eta_0$ | Under reporting factor | 1 - 10 | [7] |
| $\lambda$ | Sensitivity decay | 0 - 0.03 | (Calibrated) |
| $c$ | Cost of maintaining measures | 0 - 0.1 | [15] |
| $I_0$ | Initial infection rate | $1/\text{pop} - \alpha(1/\text{pop})$ | (Calibrated) |
| $x_0$ | Initial population support for NPIs | $0 - 1$ | (Calibrated) |
| $Ii_0$ | Initial Incidence rate | $0 - \alpha(1/\text{pop})$ | (Calibrated) |

**Table S** 2: Summary table for parameter and initial conditions ranges. $\alpha$ is a proportionality constant and "*pop*" is the population size of the respective countries understudy.

**Statistical Tools.** In this section, we will explore few statistical tools to assess the identifiability of the estimated particles with respect to the models and empirical data. We analyse our models with respect to estimated parameter values using the adjusted Akaike Information Criterion (AICc), log-likelihood, and Adjusted $R^2$ score. These enable us to assess the goodness-of-fit and compare the performance of the models. These analyses are based on the average simulation from 100 selected particles from the posterior distribution.

1. **Adjusted Akaike Information Criterion (AICc):** We assess how well the model(s) fits to the empirical data and make model comparison using AICc. We calculate AICc values using the formulae (Equation 18) since we fit the coupled model to two different empirical times series data, that is, daily reported cases for incidence and oxford stringency index for population behaviour as presented by [11] for multiple datasets.

$$\textbf{AICc} = \sum_{i=1}^{N} \Big( n_i \ln \frac{RSS}{n_i} \Big) + 2KN \Big( \frac{n_t}{n_t - KN - N} \Big) \tag{18}$$

   where $N$ is the number of data sets (N=2), $i$ is index of a data set $\{1, 2\}$ and $n_t$ is the total number of data points used in the set (i.e., in all N datasets). This equation is literally the sum of the AICc's for the two different data set, ie, AICc = $\text{AICc}_1$ + $\text{AICc}_2$. **Residual Sum of Squares (RSS)** is expressed as

$$\textbf{RSS} = \sum_{i=1}^{N} \frac{(y(\theta) - y_0)^2}{N} \tag{19}$$

   where $y(\theta)$ is the model evaluated at the particle $\theta$, $y_0$ is the empirical data and $N$ is the length of data (observation). RSS can equally be used to assess the model performance, but due to it naivety, it is not considered for direct analysis in this study.

2. **Log-likelihood:** This statistical metric is used to assess how well a model fits to empirical data and where a higher value corresponds to a better model. This enables us to also evaluate the individual

models and also compare their performance. The expression for the log-likelihood is

$$\ell(\theta) = \log L(\theta) = \sum_{i=1}^{n} \log f(x_i|\theta)$$

We assume the residuals $(e_i)$ follow a normal distribution with mean of 0 and variance of $\sigma$, thus

$$\ell(\theta) = -\frac{1}{2}\sum_{i=1}^{n} \log 2\pi\sigma^2 + \sum_{i=1}^{n} \frac{e_i}{\sigma^2} \quad (20)$$

We take the negative of this to obtain the negative log-likelihood for our analysis.

3. **Adjusted R Squared ($R^2_{adj}$)** gives the $R^2$ score, commonly referred to as the coefficient of determination, which measures the variations between the data and prediction. We use the adjusted expression since the two models have different numbers of parameters (penalising for the number of parameters) [10].

$$R^2_{adj} = 1 - \frac{\left(\frac{\mathbf{RSS}}{N-K-1}\right)}{\left(\frac{\sum(y_0-\bar{y_0})^2}{N-1}\right)}$$

### Supplementary Results

We evaluated how well the model(s) fit to the empirical data (daily reported incidences and Oxford stringency index). The estimated parameter values are expected to be meaningful and provide accurate information on the dynamics of the virus transmission. These values are considered to evaluate the predictability of both models with validation from empirical data. Since our aim for this study is to evaluate the predictiveness of our models, it is safe to consider how well both disease and coupled behaviour-disease models fit with the observation after selecting parameter values based on ABC SMC. We evaluate the performance of the model(s) by examining the posterior distribution of the estimated particles, the time series trajectory, peaks analysis, statistical measures using the adjusted $R^2$ score, an adjusted Akaike Information Criteria (AICc) for the measure of goodness of fit, and log-likelihood as discussed in previous section. We further looked at some parameter planes to check the influence of model behaviour parameters in the coupled model. For prediction analysis, we discuss the time series, peak(s), area between the model estimates and observation, and statistical measures using Adjusted AICc (see main text) and Quantile-Quantile (Q-Q) of the residuals to check nature of distribution. These analyses and observations are based on the average simulations of 100 selected particles. We note here that the empirical data used for fitting is somewhat different from data used for prediction and as such cross-validation was not considered.

We consider daily reported incidences of SARS-CoV-2 transmission in thirteen(13) European countries for our analysis before vaccine roll-out. Thus, the period of February 2020 to January 2021 is extracted for our study. Figure 1 (see main text) shows the selected countries with the daily reported cases trajectory and their corresponding stringency index as a behavioural response to the implementation of non-pharmaceutical interventions (NPIs). Clearly, we observe a sharp response from the government(population) to the reported incidences. An interesting dynamics is witnessed where a moderately high incidence is responded with very high level of stringency at the early stages (March - May). As the disease progresses to greater levels over time (September-December) we see a similar level of response by the population as before by the population as in the early stages of the outbreak for some countries, while others have reduced response (e.g., Finland, Norway).

**Model Training.** We fit both the disease model and the coupled behaviour-disease model to empirical data as discussed in the method section above. A portion of the empirical data collected for this study is used for the fitting process for each country based on the occurrence of their first major peak between March and May 2020. The length of the individual country data is different as a result of the different dynamics of the virus in the various countries. To obtain good and meaningful parameter estimates for our analysis, it was inappropriate to use the same number of data points for all countries. Table S1 shows the length of the data points used for fitting each country from the start date. The number of data points used ranges from 200 to 250 with corresponding dates February 2 to October 15, 2020.

We analyse the time-series trajectory of the model output with the empirical data (daily reported incidences) with focus on the dynamics over time. We observe that the trend of the model(s) is very similar to that of case notifications. The first major peak identified in Spring 2020 period was estimated by both models for the incidence (Fig. S 2). We also observe that the coupled behaviour-disease model also fits very well to the stringency index for the population behaviour (see main text Figure 4) except for Belarus and Finland. With respect to comparison between the disease model (SIR) and the coupled behaviour-disease model (SIRX), we note that 95% credible interval for SIRX covers most of the data points compared to the SIR model, giving us a better estimate overall in terms of trend.

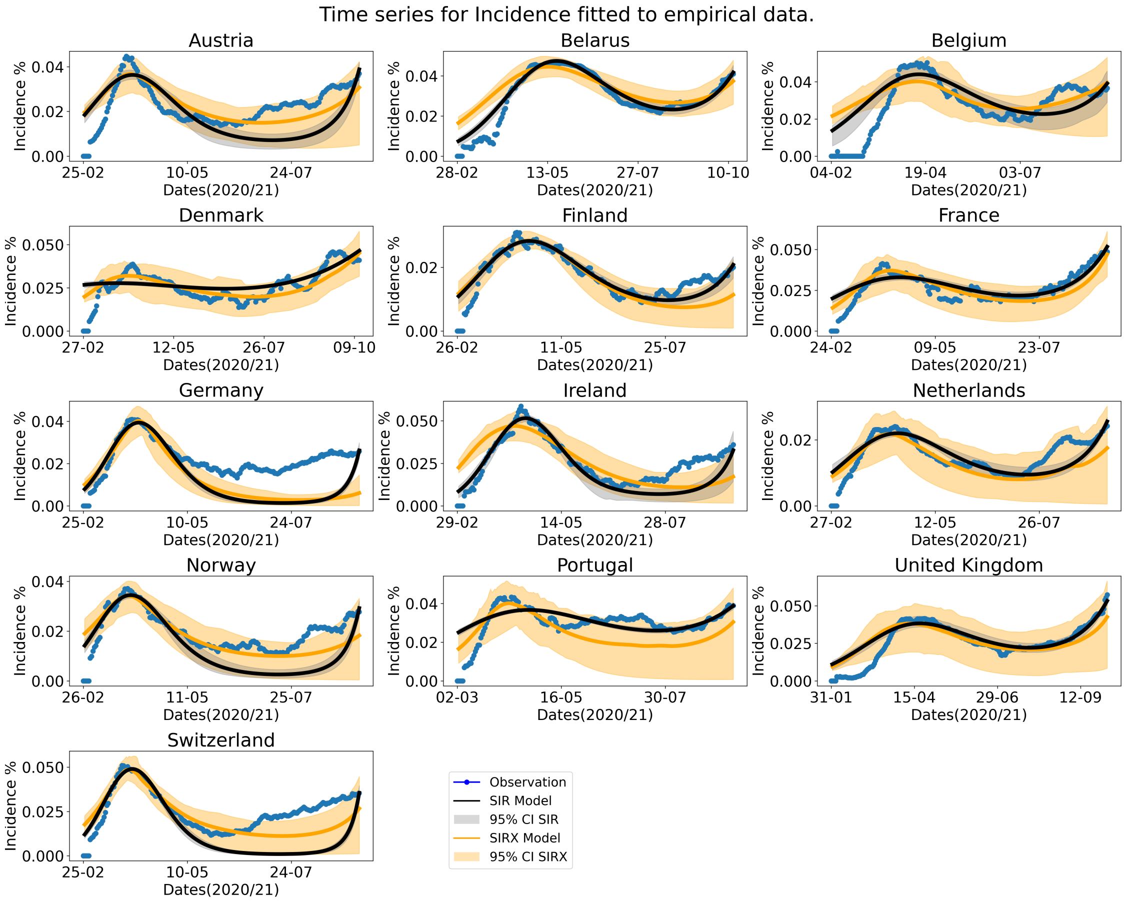


**Fig. S 2: Trajectory of disease model (SIR) and coupled behaviour-disease model (SIRX) from estimated particles from the model(s) training**. We show the actual observation, average simulation and the 95% credible interval for the fitting period.

As much as we have similar trends for both empirical data and models, we also wanted to estimate the peak values from both models using selected parameter values. We observed both models do exactly that; the estimated peak period falls within the March-May period. The average simulation predicts the peak day within a margin of error of 7 days for the two models for Austria, Belarus, Germany, Ireland, the Netherlands, Norway, and Switzerland. Additionally Denmark, France and Portugal is identified by the SIRX model to give an estimation which satisfies the 7-day error margin from that of the observation peak, while as we observe Belgium and the United Kingdom, to have good estimates for SIR model. The estimated peak period by the SIRX model for Belgium is within $(46, 97)\, 95\%CI$ and the United Kingdom within $(44, 143)\, 95\%CI$. Although the average peak for these two countries with respect to the SIRX model does not fall within our error margin, it falls within the credible interval of 95%. Fig. S (3 b) shows a summary of the actual data peak and model(s) peak period with 95% credible intervals. For the peak magnitude, we observe that the actual values are within the credible interval 95% estimated by the SIR model for Belarus, the Netherlands and the United Kingdom. The actual peak magnitude for Ireland is higher than the upper bound estimates for the two models, thus, both models are unable to estimate that for Ireland. The three countries estimated by SIR model and the remaining countries; Austria, Belgium, Denmark,Finland, France, Germany, Norway, Portugal, and Switzerland have their actual peak values within the 95% credible interval for the SIRX model. Therefore, SIRX model predicts the peak magnitude and period for at least 12 out of 13 countries base on their credible interval which is wide due to nature of coupled behaviour-disease models. Fig S (3 a) illustrates the peak magnitude identified from the actual data and the estimates made by both models.

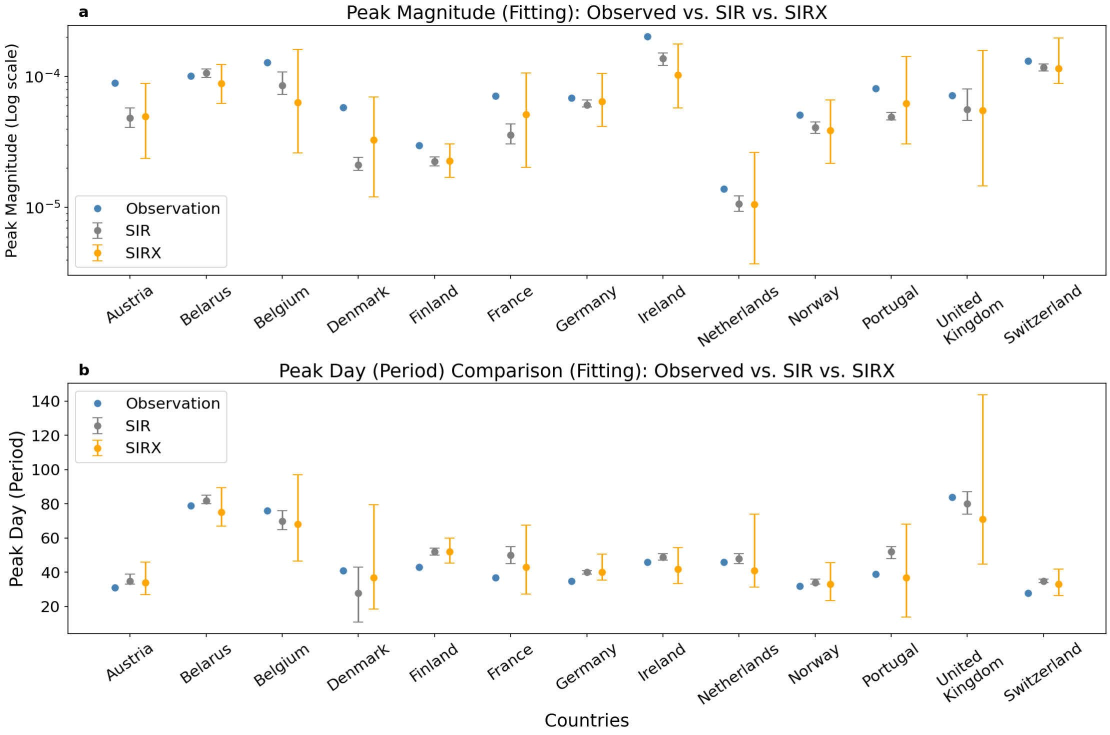


**Fig. S** 3: **Actual peak magnitude (a) and peak day (b) compared to model estimates**. We show the identified peak details and compare with estimation from disease model (SIR) and coupled behaviour-disease model (SIRX) with 95% credible interval.

As discussed in the statistical analyses, we estimate Adjusted AIC to assess the goodness-of-fit and for model comparison. Fig. S 4 illustrates the respective values for the models for 13 countries. We calculate these values for SIR incidence, SIRX incidence and SIRX incidence + behaviour. Clearly, SIRX incidence + behaviour outperform the others.

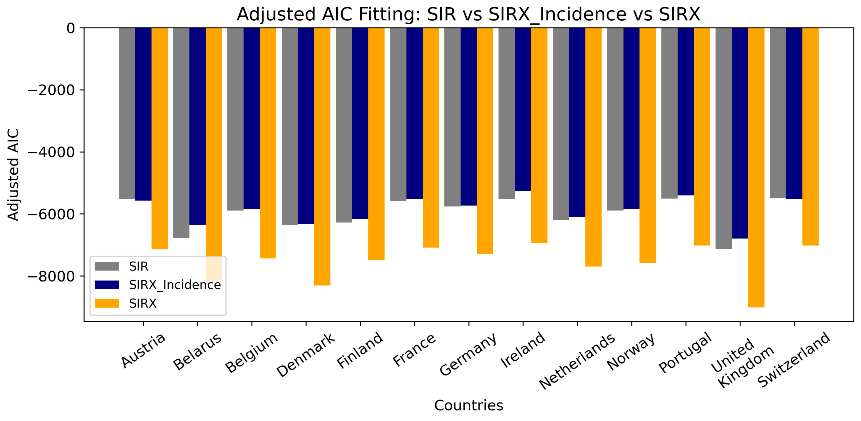


**Fig. S 4: Adjusted AIC estimates for SIR incidence, SIRX incidence, and SIRX incidence + behaviour.** Estimates are obtained using first data point to Cutoff used for fitting model to observation.

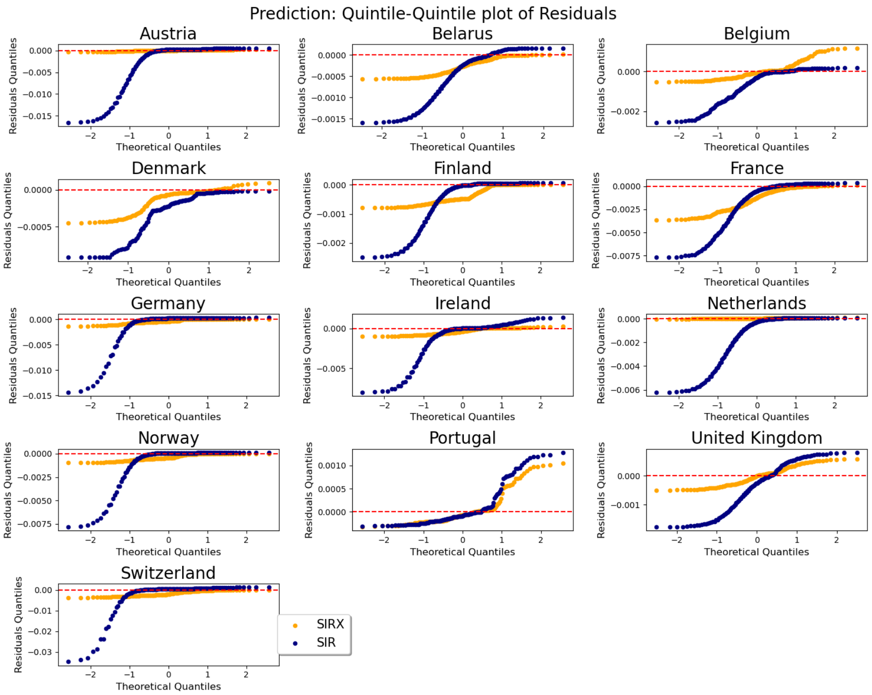


**Fig. S 5: Quantile-quantile plot of the residual assessed against a theoretical distribution (normal).** Distance between the data and model estimate as residuals against normal distribution.

The 'best' particle (with lowest MSE) posterior distribution of our parameter estimation for each iteration is illustrated in (Fig. S 6 and Fig. S 7) to show the convergence of our parameter estimation (fitting) algorithm. The sequence of errors converges as $T \to \infty$ for both models as expected. The last plot of these figures shows the lowest error for all particles for 13 countries. We see that the lowest error did not necessarily occur at the last iteration ($T$). This then implies that setting the right value for $T$ and threshold was very necessary to obtain good results for the model training for the individual countries.

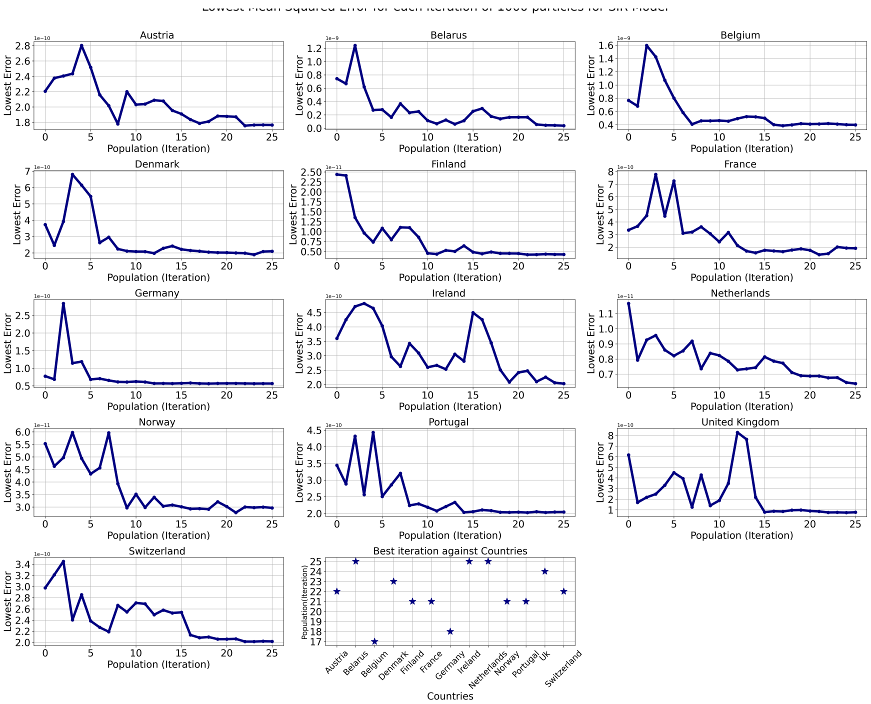


**Fig. S 6**: **Trajectory of lowest error for each iteration.** Trajectory of Lowest error (1 out 1000) of each iteration ($T = 25$) particles for the various countries. The last plot shows the iteration with the lowest error corresponding to SIR model.

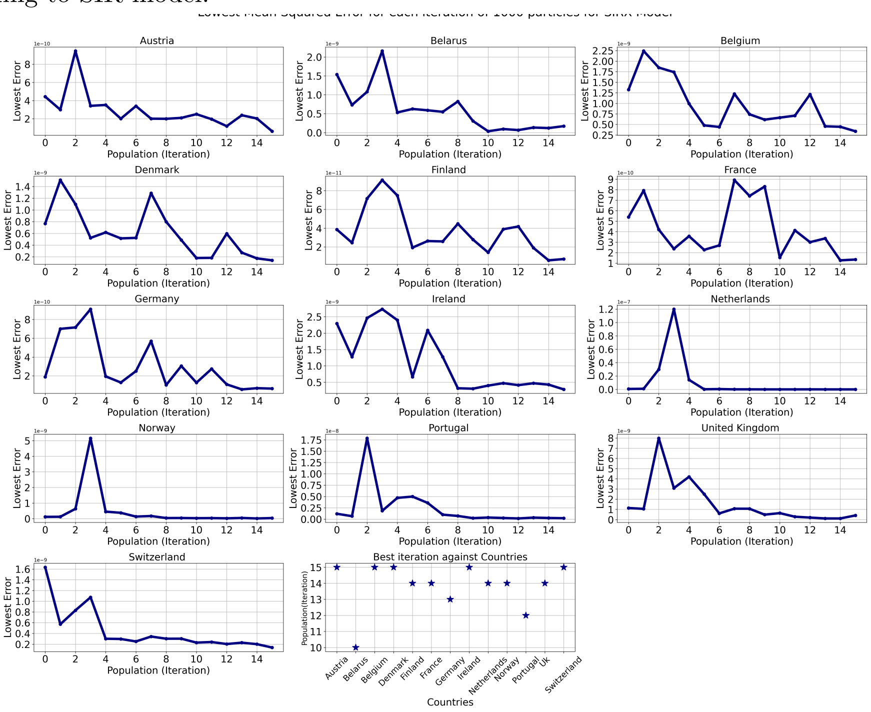


**Fig. S 7**: **Trajectory of lowest error for each iteration.** Trajectory of Lowest error (1 out 1000) of each iteration ($T = 15$) particles for the various countries. The last plot shows the iteration with the lowest error corresponding to SIRX model.

We observed that while the SIR model performs well during training, it does not perform quite well during prediction as we would expect. The SIRX model, on the other hand, performs well during fitting and also during prediction. Detailed discussions are presented in the main text on the prediction results. We investigate further the results using the basic reproduction number with seasonality ($R_0(t)$). We obtain the expression below for $R_0(t)$;

$$R_0(t) = \frac{\beta(1 + b\cos(t - \phi)}{\gamma}. \tag{21}$$

$R_0(t)$ is time dependent and takes different values at different periods of the pandemic. Fig. S 8 shows the trajectory of $R_0(t)$ for both models with 95% credible interval. We discovered that the values range from 1 to 6, consistent with other studies [1, 18]. For SIRX model $R_0(t) \in [1, 4]$ since the behaviour feedback is not factored in $R_0(t)$ estimation whereas the SIR model has $R_0(t) \in [1, 6]$. For the first 150 days which covers the Spring period while the SIR model was between 1 and 3, the SIRX model was between 1 and 2 for the selected countries. This difference is due to the term $(1 - \epsilon x)$ for the behaviour, which has influence on the transmission of the virus in the SIRX model. After 200 days, Fall period (September - December), we see that the upper bound for some countries increase significantly and a few others seem not to increase much. However, we identified five countries of interest, Austria, Ireland, the Netherlands, Norway, and Switzerland, for the SIR model. These countries have $R_0(t)$ to be as high as 6 which reflects in the predicted outcome for such model, producing dramatic prediction. The second peaks of these countries are relatively high to the empirical data due to high values for $R_0(t)$.

The basic ($R_0$) and effective($R_{eff}$) reproduction numbers are also estimated (Table S 3). The table shows the median values and 95% credible interval for coupled model without mitigation, disease model and the coupled model with mitigation as ($R_{eff}$). $R_{eff}$ from the coupled model is lower compared to $R_0$ for the disease model. This shows the effect of mitigation to reduce transmission as is the goal of mitigators. The median and upper bound values for France for the coupled model is higher than estimates from the disease model, every other country has the opposite as the case.

**Table S3. Estimation of basic ($R_0$) and effective ($R_{eff}$) reproduction numbers.**

| Countries | SIRX ($R_0$) | SIR ($R_0$) | $R_{eff}$ |
|---|---|---|---|
| Austria | $1.333 \in (0.786, 1.566)$ | $2.905 \in (0.243, 3.598)$ | $1.321 \in (0.678, 1.545)$ |
| Belarus | $1.409 \in (0.835, 1.796)$ | $1.631 \in (1.16, 1.76)$ | $1.405 \in (0.835, 1.788)$ |
| Belgium | $1.43 \in (0.938, 1.786)$ | $2.159 \in (0.837, 2.699)$ | $1.313 \in (0.541, 1.626)$ |
| Denmark | $1.638 \in (0.958, 2.361)$ | $1.536 \in (1.173, 1.664)$ | $1.553 \in (0.614, 2.159)$ |
| Finland | $1.492 \in (0.493, 1.819)$ | $1.611 \in (1.057, 1.766)$ | $1.461 \in (0.493, 1.783)$ |
| France | $2.02 \in (0.702, 2.996)$ | $1.515 \in (1.157, 1.6)$ | $1.908 \in (0.454, 2.558)$ |
| Germany | $2.331 \in (0.145, 2.964)$ | $2.264 \in (0.684, 2.562)$ | $2.306 \in (0.067, 2.931)$ |
| Ireland | $2.027 \in (0.141, 2.476)$ | $3.584 \in (0.024, 4.348)$ | $1.991 \in (0.131, 2.424)$ |
| Netherlands | $1.823 \in (1.127, 2.636)$ | $2.424 \in (0.644, 2.931)$ | $1.512 \in (0.426, 2.106)$ |
| Norway | $1.288 \in (0.705, 1.483)$ | $2.481 \in (0.418, 3.16)$ | $1.26 \in (0.632, 1.438)$ |
| Portugal | $1.882 \in (0.621, 2.805)$ | $1.418 \in (1.217, 1.455)$ | $1.809 \in (0.339, 2.703)$ |
| United Kingdom | $1.548 \in (0.769, 1.998)$ | $1.534 \in (1.152, 1.654)$ | $1.537 \in (0.677, 1.972)$ |
| Switzerland | $2.148 \in (0.141, 2.624)$ | $2.759 \in (0.344, 3.372)$ | $2.067 \in (0.107, 2.537)$ |

**Table S** 3: Estimation of basic ($R_0$) and effective ($R_{eff}$) reproduction numbers for the prediction period – after 200 days.

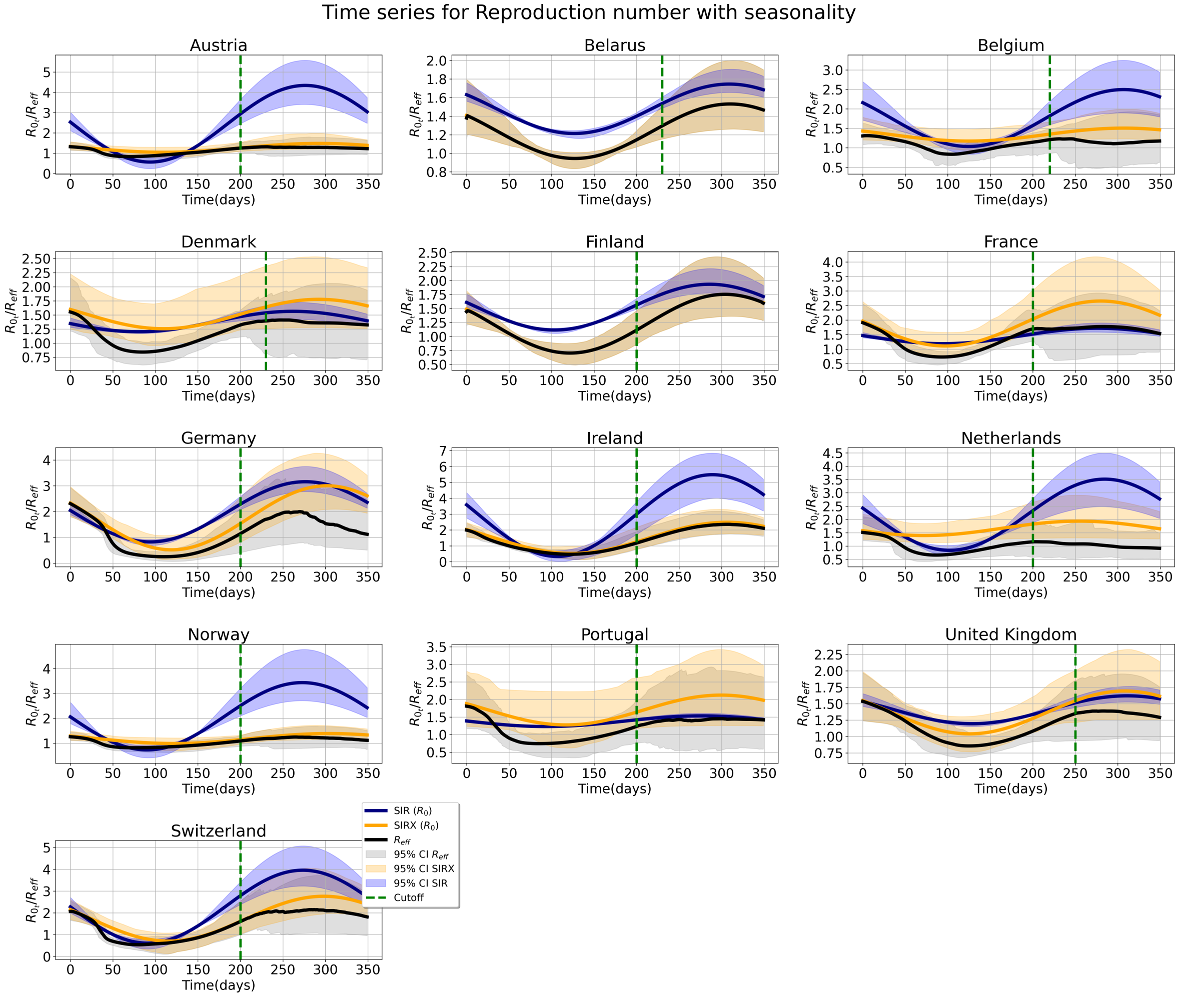


**Fig. S** 8: **Basic ($R_0$) and Effective ($R_{eff}$) reproduction numbers**. $R0$ and $R_{eff}$ as time dependent for all 13 countries for both models with 95% credible intervals.

**Parameter Planes.** We evaluate the influence of the parameters; $\epsilon, \lambda, c, \kappa$ and $x_0$ which are attributed to the coupled behaviour-disease model. The parameter plane looks at the effects of different combinations of two parameter values on the peak magnitude and period.

**Definition of Peak:** Peak is defined to be the maximum value within a given period. For empirical data, it is the maximum number of cases reported by each country and for the models, it is the maximum point in the average simulation. The value becomes the peak magnitude and the index becomes the peak period (day).

Using the particle with lowest error, we substitute different values for the behaviour parameters to assess their influence on peak values (magnitude and period). We do this for the fitting and the prediction separately. We observe some form of influence of the parameters on the peaks where we identified some critical transition and others to have stable regions. The stable regions imply the peak is not affected significantly by a change in the values of such parameters. While some have sharp transitions, others have rough transitions and no clear pattern.

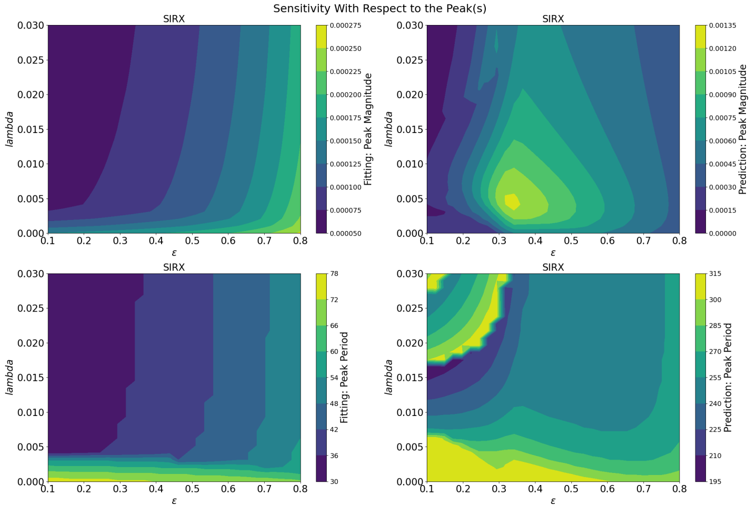


**Fig. S 9: Sensitivity analysis examining a range of values for sensitivity decay and intervention effectiveness on peak prediction.** Influence of $\lambda$ and $\epsilon$ on the peak magnitude and period for the first peak and second peak respectively estimated by the SIRX model.

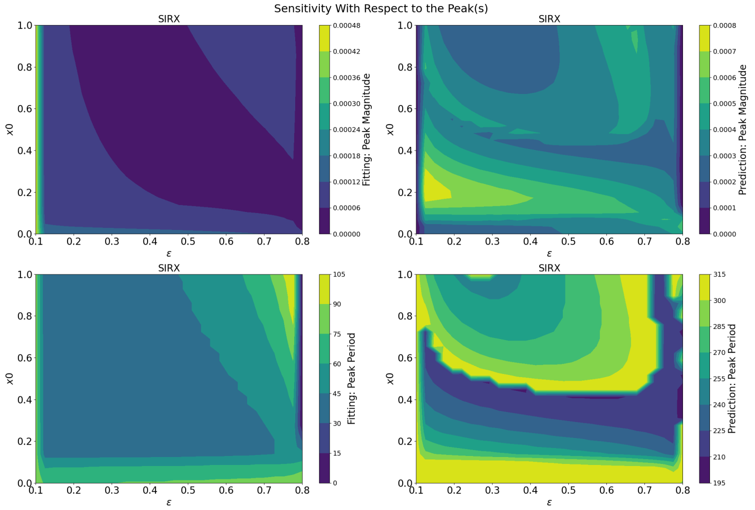


**Fig. S 10: Sensitivity analysis examining a range of values for initial mitigation support and intervention effectiveness on peak prediction.** Influence of $x_0$ and $\epsilon$ on the peak magnitude and period for the first peak and second peak respectively estimated by the SIRX model.

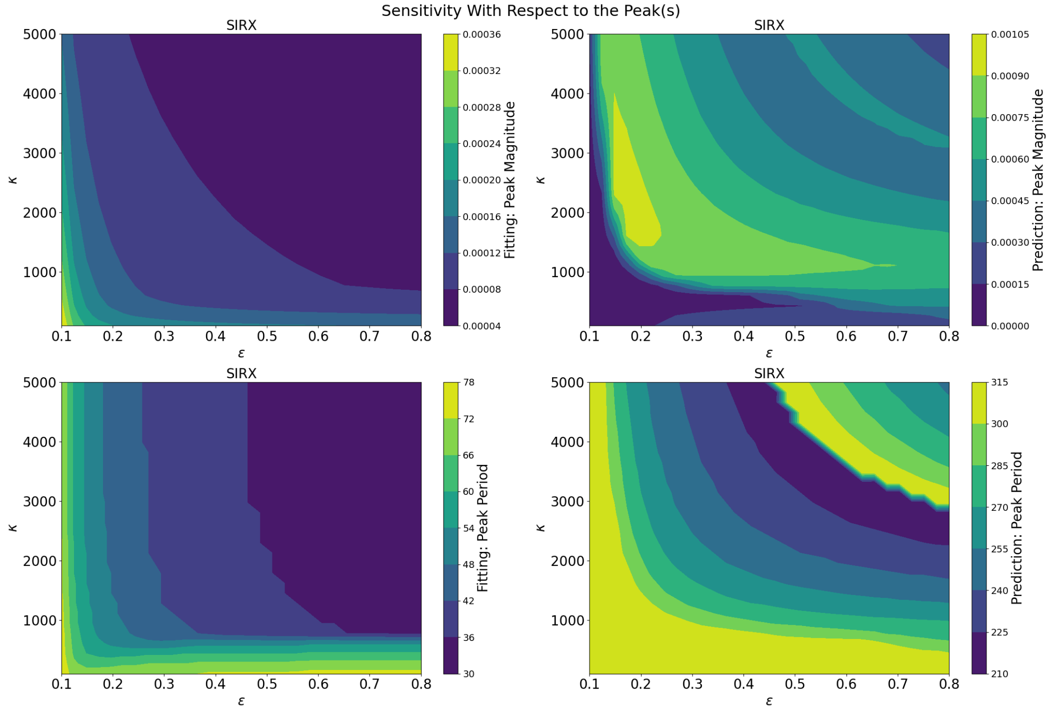


**Fig. S** 11: **Sensitivity analysis examining a range of values for social learning rate and intervention effectiveness on peak prediction.**Effect of $\kappa$ and $\epsilon$ on the peak magnitude and period for the first peak and second peak respectively estimated by the SIRX model.

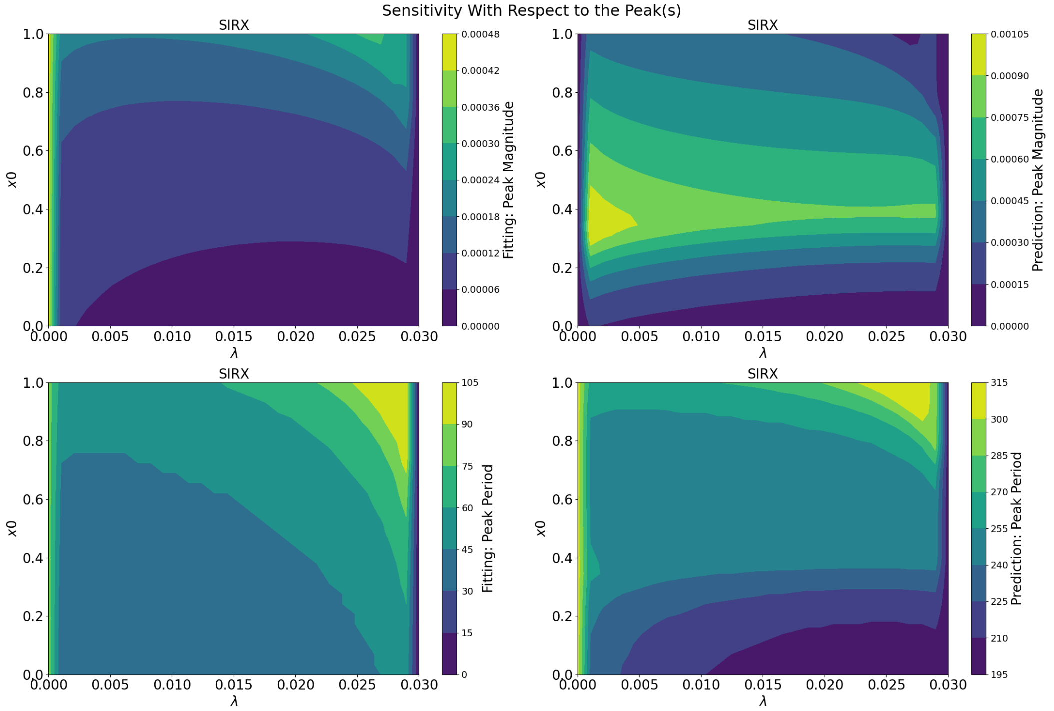


**Fig. S** 12: **Sensitivity analysis examining a range of values for initial mitigation support and sensitivity decay on peak prediction.** Effect of $x_0$ and $\lambda$ on the peak magnitude and period for the first peak and second peak respectively estimated by the SIRX model.

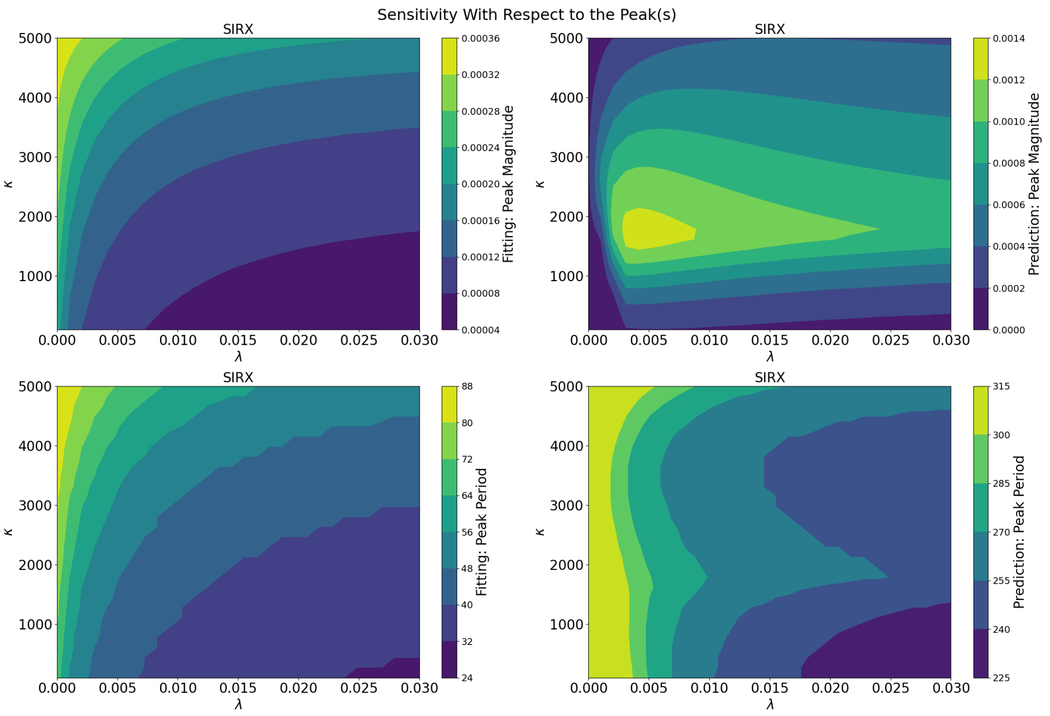


**Fig. S 13: Sensitivity analysis examining a range of values for social learning rate and sensitivity decay on peak prediction.** Effect of $\kappa$ and $\lambda$ on the peak magnitude and period for the first peak and second peak respectively estimated by the SIRX model.

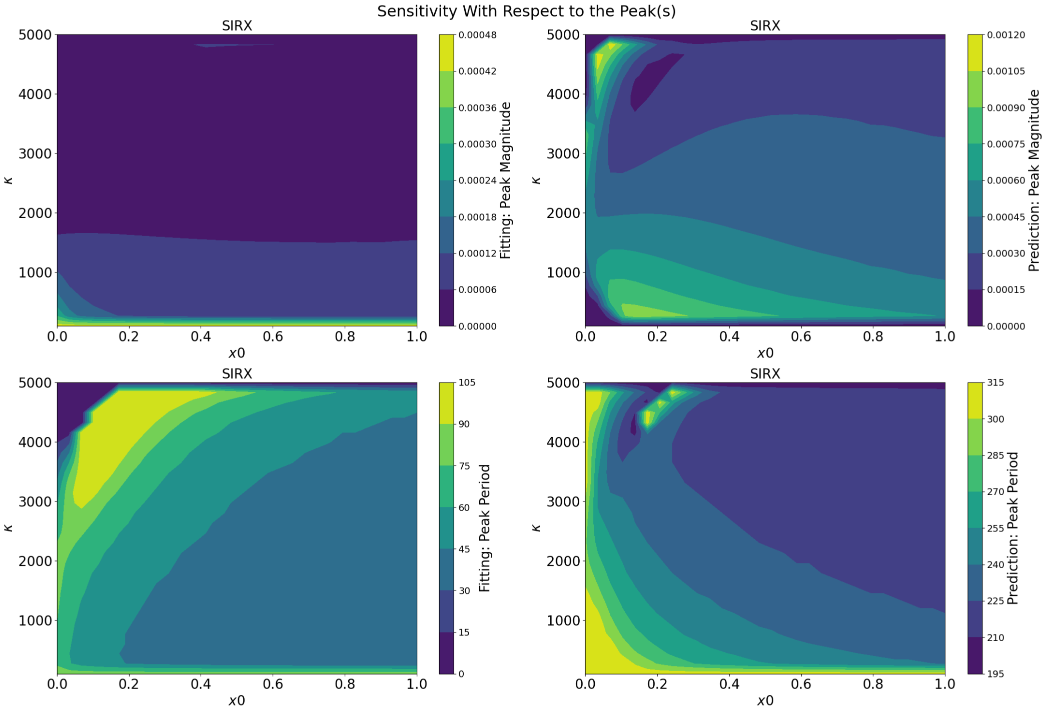


**Fig. S 14: Sensitivity analysis examining a range of values for social learning rate and initial mitigation support on peak prediction.**Effect of $\kappa$ and $x_0$ on the peak magnitude and period for the first peak and second peak respectively estimated by the SIRX model.

**Particle Analysis.** Parameter estimation as discussed in the methods section uses ABC SMC from which we select particles with lowest error (MSE) values. For our analysis, we used 100 out of 1000 particles per iteration from the posterior distribution ($\geq$ 15000 particles) obtained for each model and for each individual country, respectively. We observe that the posterior distribution for parameters for the disease model (SIR) is clustered to a section of the initial parameter range values unlike the coupled model which is spread across (see Fig. S 15 for Austria). The posterior distribution of estimated particles for both models are either symmetric or asymmetric, that is, unimodal, bimodel, left or right skewed (Fig. S 16) in the case for Austria. For SIR model the distribution for the phase of seasonality ($\phi$) is uni-modal while as the other parameters take different forms of distributions. The phase of seasonality for the SIRX model is left skewed but transmission rate ($\beta$) is uni-model. We observed similar results for the other countries and no major differences from what has been discussed. From Fig S 15, we observe these selected particles for both models have relatively very low errors. However, we note that the disease model (SIR) particles estimates on average has lower MSE value than the coupled model using the 6 common parameters. This could imply a better estimate for the SIR model compared with the SIRX model (see Fig. S 2). Due to the clustered nature of the SIR model estimate, we observe the range for the 95% credible interval is narrower compared to SIRX model which has a wide range. This case for Austria is very similar to the cases of the 12 other countries.

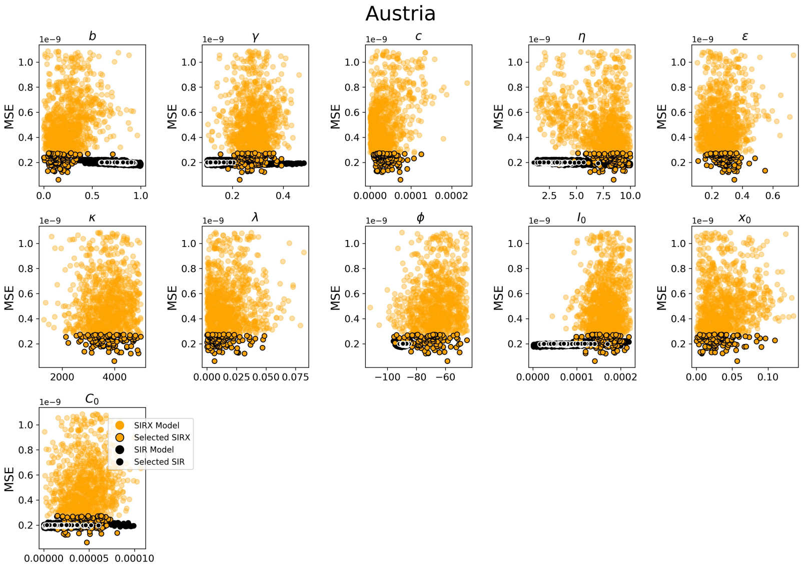


**Fig. S** 15: **Posterior distributions on the inferred parameters for both models against MSE.** Distribution of 1000 particles estimated for the parameter set for both disease (SIR/black) and coupled behaviour-disease (SIRX/orange) models. The values are plotted against their mean squared error (MSE) and are the output for the "best" iteration (population) in our ABC SMC approach for parameter estimation.

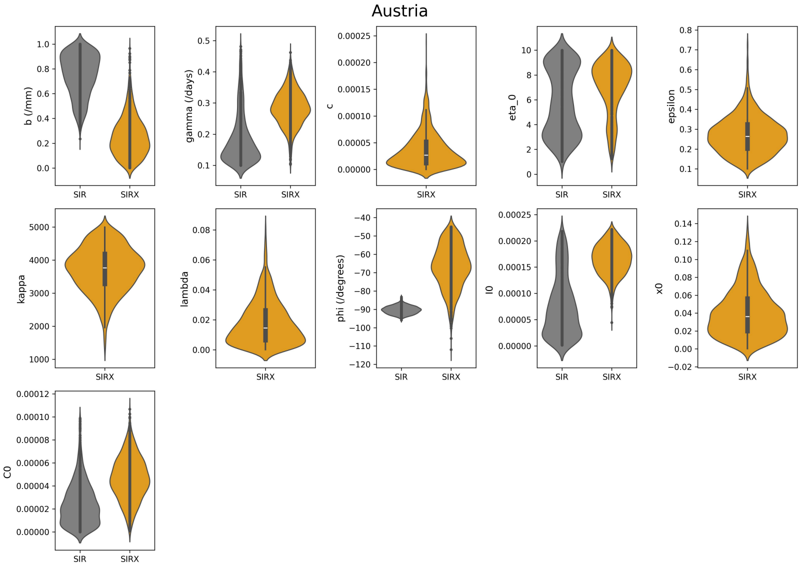


**Fig. S** 16: **Posterior distributions on the inferred parameters for both models.** Distribution of 1000 particles estimated for the parameter set for both disease (SIR/black) and coupled behaviour-disease (SIRX/orange) models. This is illustrated using a violin plot which enables us to assess the probability density function of the posterior distribution.

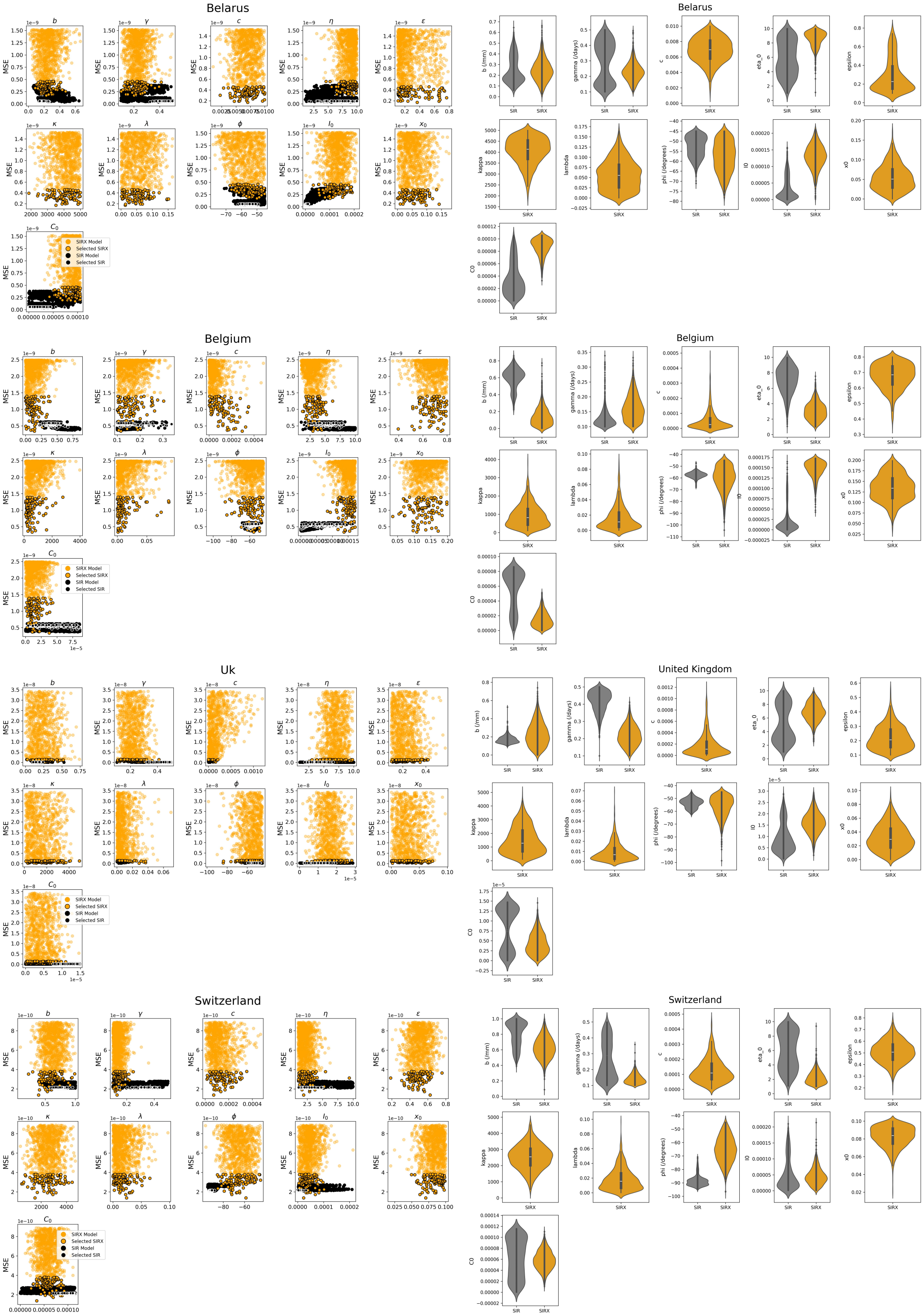


**Fig. S 17**: **Posterior distributions on the inferred parameters for both models.** Distribution of parameters against the mean squared error of 1000 particles and selected 100 particles used for the analysis. And a violin plot (right figure) which shows the type of distribution of the particles.

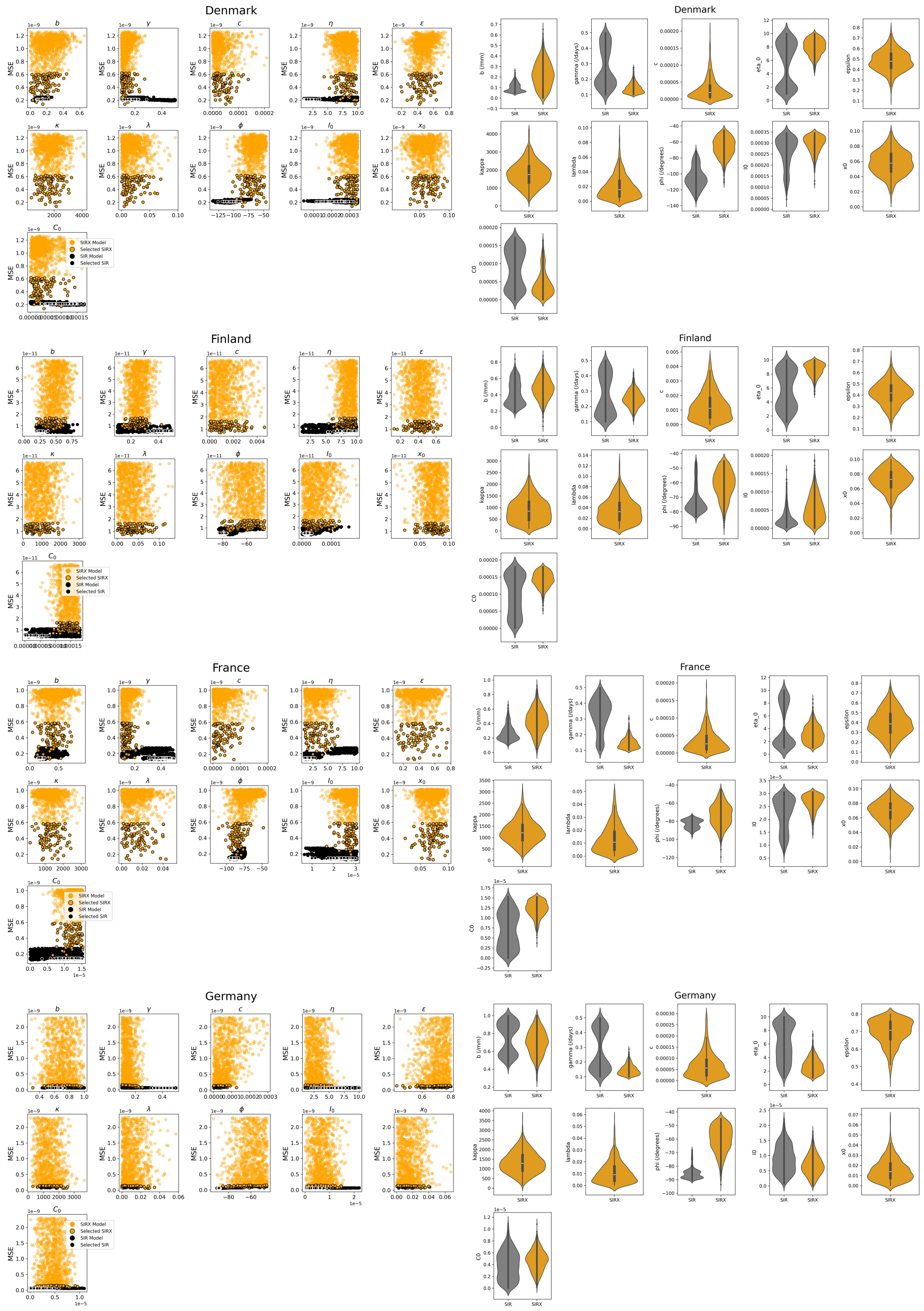


**Fig. S** 18: **Posterior distributions on the inferred parameters for both models.** Distribution of parameters against the mean squared error of 1000 particles and selected 100 particles used for the analysis. And a violin plot (right figure) which shows the type of distribution of the particles.

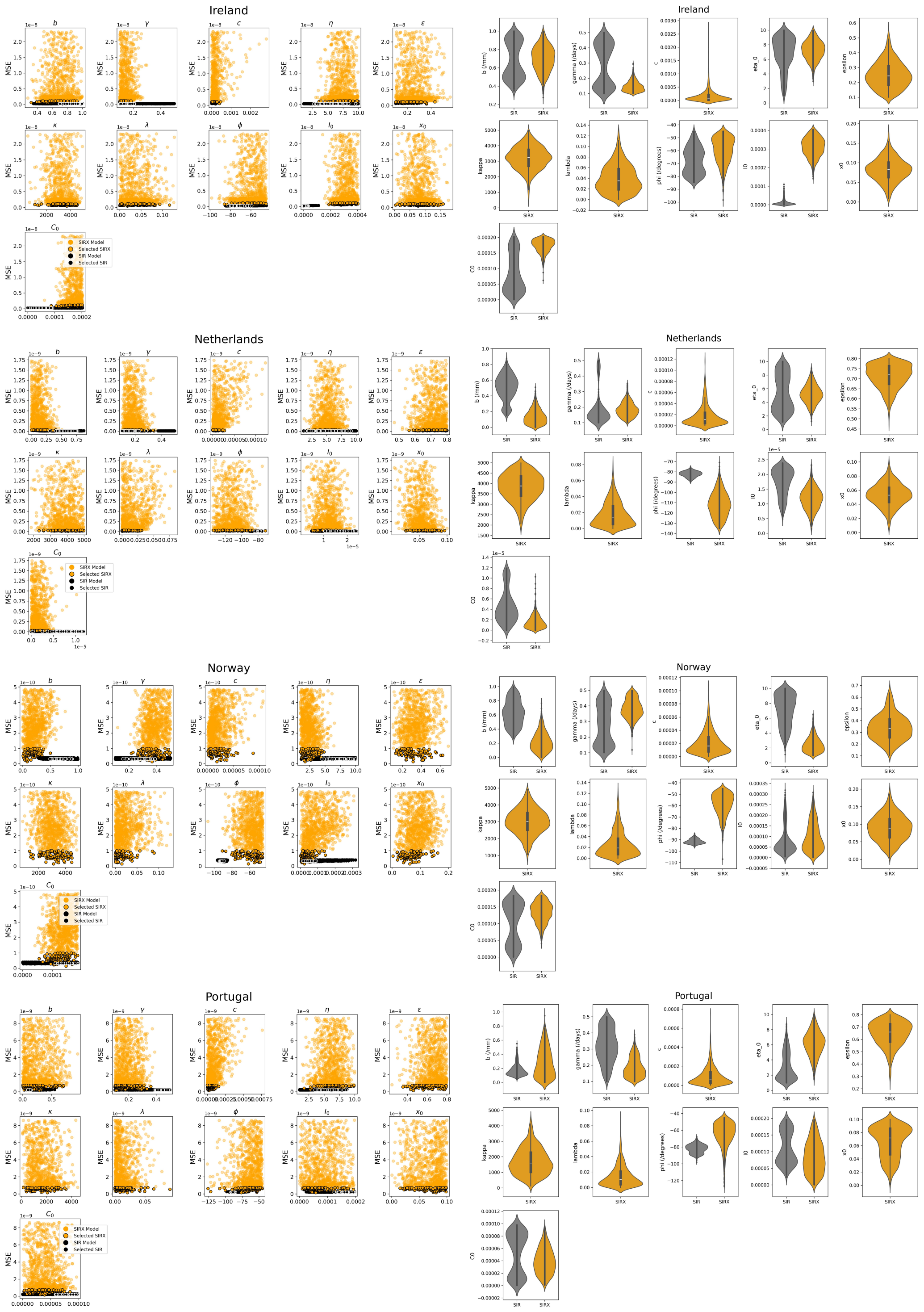


**Fig. S** 19: **Posterior distributions on the inferred parameters for both models.** Distribution of parameters against the mean squared error of 1000 particles and selected 100 particles used for the analysis. And a violin plot (right figure) which shows the type of distribution of the particles.

**Table S4. Summary of Statistical metrics for 13 countries after Training models on test data.**

| Countries | Model | Neg Log-Likelihood | $R^2_{adj}$ | AICc |
|---|---|---|---|---|
| Austria | SIRX Model Incidence | 92.3 | 0.999946 | -5569.72 |
| | SIRX Model Behaviour | 152.76 | 0.991947 | -1571.68 |
| | SIR Model | 98.07 | 0.999929 | -5522.45 |
| Belarus | SIRX Model Incidence | 108.34 | 0.99962 | -6353.17 |
| | SIRX Model Behaviour | 939.36 | 0.965996 | -1797.52 |
| | SIR Model | 104.82 | 0.999939 | -6777.57 |
| Belgium | SIRX Model Incidence | 101.81 | 0.999848 | -5834.46 |
| | SIRX Model Behaviour | 433.56 | 0.988405 | -1596.38 |
| | SIR Model | 100.18 | 0.999882 | -5896.74 |
| Denmark | SIRX Model Incidence | 113.91 | 0.999825 | -6313.2 |
| | SIRX Model Behaviour | 183.34 | 0.991067 | -1984.57 |
| | SIR Model | 104.91 | 0.999855 | -6362.77 |
| Finland | SIRX Model Incidence | 99.78 | 0.999872 | -6166.75 |
| | SIRX Model Behaviour | 733.47 | 0.960174 | -1315.45 |
| | SIR Model | 88.59 | 0.999926 | -6281.13 |
| France | SIRX Model Incidence | 99.76 | 0.999934 | -5516.5 |
| | SIRX Model Behaviour | 112.56 | 0.991493 | -1571.47 |
| | SIR Model | 89.85 | 0.999953 | -5592.96 |
| Germany | SIRX Model Incidence | 189.23 | 0.999915 | -5728.28 |
| | SIRX Model Behaviour | 113.58 | 0.985959 | -1573.63 |
| | SIR Model | 144.03 | 0.999924 | -5758.31 |
| Ireland | SIRX Model Incidence | 91.1 | 0.999593 | -5260.1 |
| | SIRX Model Behaviour | 204.54 | 0.99651 | -1683.40 |
| | SIR Model | 95.06 | 0.999881 | -5512.11 |
| Netherlands | SIRX Model Incidence | 144.73 | 0.999943 | -6111.58 |
| | SIRX Model Behaviour | 107.07 | 0.991733 | -1581.82 |
| | SIR Model | 92.33 | 0.999961 | -6191.14 |
| Norway | SIRX Model Incidence | 104.0 | 0.999713 | -5841.16 |
| | SIRX Model Behaviour | 143.5 | 0.995688 | -1744.60 |
| | SIR Model | 119.2 | 0.999768 | -5888.82 |
| Portugal | SIRX Model Incidence | 195.98 | 0.999935 | -5406.98 |
| | SIRX Model Behaviour | 148.08 | 0.989078 | -1612.97 |
| | SIR Model | 89.98 | 0.99996 | -5510.44 |
| United Kingdom | SIRX Model Incidence | 126.63 | 0.999901 | -6789.74 |
| | SIRX Model Behaviour | 173.72 | 0.99786 | -2218.5 |
| | SIR Model | 115.28 | 0.999974 | -7133.65 |
| Switzerland | SIRX Model Incidence | 113.45 | 0.999978 | -5515.06 |
| | SIRX Model Behaviour | 150.38 | 0.982795 | -1507.46 |
| | SIR Model | 125.22 | 0.999976 | -5498.57 |

**Table S** 4: Summary values for negative log-likelihood, adjusted $R^2$ and adjusted AIC for both models.

Our results show that both models fit pretty well with our empirical data; however, due to the fact that not one particular model was identified by all 3 metrics to be the better model for all 13 countries, we will not attempt to generalise that either SIR model or SIRX model is better model in terms of model fitting (training). In light of that, we note SIR model gives a good average compared to the SIRX model but a narrow interval for the 95% credible interval. The SIRX model on the other hand, gives a wide interval for the 95% credible interval which contains most of the empirical data due to the nature of social-disease models. This difference in the credible interval can be attributed to the posterior distribution of the estimated particles for both models.
